# The Role of Age and Sex in Non-linear Dilution Adjustment of Spot Urine Arsenic

**DOI:** 10.1101/2023.04.12.23288341

**Authors:** Thomas Clemens Carmine

## Abstract

**Background:** Previous research introduced V-PFCRC as an effective spot urinary dilution adjustment method for various metal analytes, including the major environmental toxin arsenic. V-PFCRC normalizes analytes to 1 g/L creatinine (CRN) by adopting more advanced power-functional corrective equations accounting for variation in exposure level. This study expands on previous work by examining the impacts of age and sex on corrective functions.

**Methods:** Literature review of the effects of sex and age on urinary dilution and the excretion of CRN and arsenic. Data analysis included a Data Set 1 of 5,752 urine samples and a partly overlapping Data Set 2 of 1,154 combined EDTA blood and urine samples. Both sets were classified into age bands, and the means, medians, and interquartile ranges for CRN and TWuAs in uncorrected (UC), conventionally CRN-corrected (CCRC), simple power-functional (S-PFCRC), sex-aggregated (V-PFCRC SA), and sex-differentiated V-PFCRC SD modes were compared. Correlation analyses assessed residual relationships between CRN, TWuAs, and age. V-PFCRC functions were compared across three numerically similar age groups and both sexes. The efficacy of systemic dilution adjustment error compensation was evaluated through power-functional regression analysis of residual CRN and the association between arsenic in blood and all tested urinary result modes.

**Results:** Significant sex differences in UC and blood were neutralized by CCRC and reduced by V-PFCRC. Age showed a positive association with blood arsenic and TWuAs in all result modes, indicating factual increments in exposure. Sex-differentiated V-PFCRC best matched the sex-age kinetics of blood arsenic. V-PFCRC formulas varied by sex and age and appeared to reflect urinary osmolality sex-age-kinetics reported in previous research.

V-PFCRC minimized residual biases of CRN on TWuAs across all age groups and sexes, demonstrating improved standardization efficacy compared to UC and CCRC arsenic.

**Interpretation:** Sex differences in UC and CCRC arsenic are primarily attributable to urinary dilution and are effectively compensated by V-PFCRC. While the sex and age influence on V-PFCRC formulas align with sex- and age-specific urinary osmolality and assumed baseline vasopressor activities, their impact on correction validity for entire collectives is minimal.

**Conclusion:** The V-PFCRC method offers a robust correction for urinary arsenic dilution, significantly reducing systemic dilution adjustment errors. Its application in various demographic contexts enhances the accuracy of urinary biomarker assessments, benefiting clinical and epidemiological research. V-PFCRC effectively compensates for sex differences in urinary arsenic. Age-related increases in TWuAs are exposure-related and should be additionally accounted for by algebraic normalization, covariate models, or standard range adjustments.

## 1. Introduction

### 1.1. Variable Power Functional Creatinine Correction (V-PFCRC)

Urinary biomarkers are essential for medical, epidemiological, and environmental research. However, variations in hydration and urine dilution can significantly affect these biomarkers. Traditional dilution adjustment methods that relate biomarker concentrations to creatinine (CCRC), osmolality, or specific gravity have limitations due to confounders such as age, sex, and muscle mass, as well as complex biochemical interactions.^1–3^

One significant limitation of the creatinine (CRN) method is that the excretion of creatinine varies with diuresis in a manner different from most analytes. This discrepancy can lead to misinterpretations, particularly at the extremes of dilution, necessitating restrictions to the accepted dilution range.^2,3^

Previous studies on dilution adjustment of arsenic and various metals demonstrated substantial neutralization of these errors by power functional modifications of the traditional ratio formations between analytes and their correctors.^2^ This led to the development of the Variable Power-Functional Creatinine Correction (V-PFCRC) method, which, in contrast to previous power-functional adjustments, further accounts for variations in corrective exponents with analyte levels.^3^ It standardizes urinary analytes to 1 g/L CRN by combining uncorrected analyte concentrations and two analyte-specific coefficients c and d according to the general V-PFCRC formula:

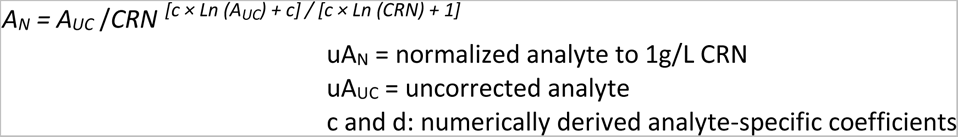

The innovative V-PFCRC formula has shown promising spot urinary adjustment results for the top priority environmental toxin arsenic, four other metals (cesium, molybdenum, strontium, zinc), and iodine. It effectively neutralized residual bias and improved correlations between blood and urine findings, indicating its potential for medical, forensic, and environmental applications.

Beyond the mathematically tangible systemic dilution adjustment error caused by differential diuresis dependencies, there are further non-linear, exposure-differentiated skews addressed by V-PFCRC. These include complex metabolic and excretion-related interactions between analytes and CRN. For instance, arsenic may limit CRN synthesis, and both may interact through the methyl donor S-Adenosyl Methionine (SAMe).^4^ Additionally, common renal tubular transport mechanisms for CRN and arsenic, such as Multidrug and Toxin Extrusion protein 1 (MATE1) or P-glycoprotein 1/Multidrug Resistance Protein 1 (PGP1/ MDR1), add to the complexity, underscoring the need for innovative approaches to address these challenges.^3–6^

While the V-PFCRC method can minimize these non-linear errors, other confounders like age, sex, and lean muscle mass, which V-PFCRC does not compensate, remain and can still influence concentrations of correctors and analytes.^2,3^ The main goal of this study was to assess whether and how far additional adjustments based on age and sex are necessary for V-PFCRC formulas or reference ranges for urinary arsenic. To accomplish this objective, the following steps were undertaken:

1. **Literature Review**: Existing literature was reviewed to understand the extent of age- and sex-specific variability in total weight arsenic urine samples and their physiological, pathological, and hormonal backgrounds.
2. **Influence of Age and Sex**: The influence of age and sex on various result modes of arsenic spot urine samples was investigated on a large dataset (Set 1) comprising 5,725 samples previously utilized to develop V-PFCRC formulas.
3. **Total Weight Arsenic Analysis in Blood and Urine**: The effects of sex and age on total weight arsenic in 1,154 urine and parallel blood samples (Set 2), substantially overlapping with Set 1, were studied.
4. **Quartile Comparison**: The blood quartile limits (1-3) and different urinary result modes were compared to assess the alignment of gender ratios and age-related changes relative to younger individuals.
5. **V-PFCRC Formula Assessment**: The age- and sex-specific influences on the coefficients of the V-PFCRC formulas were explored to evaluate the need for potential modifications.
6. **Validation**: The study further validated the variable power-functional CRN dilution correction determined in previous work by comparing residual CRN-biases of different result modes within age- and sex-specific subgroups. This rigorous validation process ensures the reliability and accuracy of the novel V-PFCRC method.

Aging and sex influence fluid balance and the metabolism of creatinine, toxins, and their respective urinary biomarkers through interconnected structural and functional changes in the kidneys and other organs. Understanding these factors is crucial for accurately adjusting spot urine samples for dilution, ensuring reliable and meaningful exposure estimates.

### 1.2. Sex Differences in Osmoregulation, Urinary Dilution, and Creatinine

Sex differences in urinary dilution and their hormonal-physiological backgrounds are summarized and referenced in Supplements 1–7. Women tend to produce higher weight-adjusted 24-hour urine volumes and display higher water turnover and natriuresis (sodium excretion).^7–9^ They typically have lower urinary concentration, osmolality (uOsm), total excreted osmotic load, and creatinine (CRN) levels than men.^7,10–12^

Sex hormones influence CRN in several ways. Androgens (male hormones) increase lean muscle mass and energy metabolism, producing nitrogenous waste products like urea, uric acid, and CRN.^13^ Men compensate for the excess CRN production mainly through increased glomerular filtration, proportional to plasma concentrations of freely filtrable CRN and renal plasma flow. Additionally, androgens enhance tubular secretion of CRN by upregulating its primary transporters, Organic Cation Transporter 2 (OCT2), for its basolateral uptake and MATE1 for its apical excretion, while estrogens inhibit these transporters.^5,10,14–16^ Male hormones also enhance the actions of the Renin-Angiotensin-Aldosterone System (RAAS) and Arginine-Vasopressin (AVP), both crucial for osmoregulation.^7,8,10,17–25^ Androgens further increase the sensitivity of AVP-immunoreactive neurons and the number of renal AVP-binding sites.^26,27^ This promotes water and sodium reabsorption, resulting in higher general urinary concentration and CRN levels.^7,10^ Conversely, female hormones inhibit these effects, either directly on the hypothalamic-pituitary axis and kidneys or through the stimulation of renal prostaglandins and NT-proBNP, which act as natural antagonists to the renal fluid-retaining actions of AVP.^20,25,28,29^ This explains the higher water turnover,^8^ natriuresis,^30,31^ urinary dilution,^7,14^ and lower extracellular volumes of women.^19,20,32,33^

### 1.3. Sex Differences in Renal Arsenic Excretion

Several studies have underscored the superior methylation capacity in women, a critical factor in the metabolic biotransformation of arsenic.^34–36^ This finding suggests that sex differences in arsenic metabolism may contribute to the generally lower female arsenic toxicity. The process involves a complex interplay of hormonal, enzymatic, and renal tubular transport factors. Women tend to dispose of higher levels of S-adenosylmethionine (SAMe), the essential methyl donor in the oxidative methylation of arsenic. This may be partially due to estrogen-dependent upregulation of choline synthesis, a critical cofactor in SAMe production,^35^ besides the lower female lean muscle mass implicating reduced SAMe consumption for creatinine synthesis. Furthermore, women exhibit higher expression of antioxidative protective enzymes such as Endothelial Nitric Oxide Synthase (eNOS), Metallothionein, Manganese Superoxide Dismutase (MnSOD), and Carbonic Anhydrase, which may further enhance the oxidative methylation of inorganic arsenic.^10,37,38^ Consistently, several sex-comparative studies reported significantly higher urinary proportions of methylated dimethylarsinic acid (%DMA) with lower inorganic arsenic (%iAs) and monomethylarsonic acid (%MMA) in women.^34,36,39–43^ In contrast to the evident sex-differentiate proportions of iAs, MMA, and DMA,^44^ no significant differences in covariate dilution-adjusted total weight arsenic and non-toxic arsenobetaine (‘fish-arsenic’) between males and females were found in a recent sizeable Korean study.^34^

The complexity of sex differences in arsenic excretion is highlighted by the male-dominant baseline expression of sodium-coupled phosphate cotransporter II (NaPi-IIa). This transporter primarily facilitates sodium reabsorption in the proximal tubule using phosphate (PO_4_^−^) as the primary co-transported anion, with inorganic arsenate (iAs_V_) able to substitute for PO_4_^−^.^34,45^ However, the cellular tubular processing of these anions differs. Plasma PO_4_^−^ levels increase with NaPi-IIa upregulation, indicating excess PO_4_^−^ reabsorption.^46^ In contrast, arsenate is partly converted to arsenite (iAs_III_) within the tubule cells, which is then either excreted in the urine or reabsorbed into the blood.^47,48^ Trivalent arsenite undergoes more efficient methylation than pentavalent arsenate in liver and kidney cells.^47,49–52^ The increased uptake of arsenate and its subsequent conversion to arsenite may lead to enhanced net methylation of excreted arsenic. This is supported by the observation of higher urinary %DMA levels in postmenopausal women, older individuals, and people with chronic kidney disease, all of whom show increased NaPi-IIa activity due to reduced expression of the proteohormone transmembrane Klotho (m-Kl).^4,39,53–61^ Estrogen inhibits NaPi-IIa transporters via upregulation of m-Kl, thus enhancing the excretion of PO_4_^−^ and iAsV.^62,63^ However, the expected sex difference in NaPi-IIa activity does not reflect in lower female serum PO_4_^−^ levels in younger adults.^53,54^ Only after menopausal estrogen decline does serum PO_4_^−^ become female-dominant. Testosterone and other factors could play more age-constant roles in inhibiting PO_4_^−^ reabsorption in males or counterbalancing the estrogen effects in females.^64^ Postmenopausal changes, including a sharper estrogen decline in women compared to testosterone in men in addition to the above-mentioned female methylation advantages, may explain higher serum phosphate and %DMA, along with lower %iAs in women.^53,54^

### 1.4. Influence of Age on Renal Function and Fluid Balance

Aging induces complex changes in renal function and fluid balance due to an interrelated combination of normal organ aging, toxic build-up, and degenerative diseases like diabetes, hypertension, atherosclerosis, and congestive heart disease.^65–68^ The combination of these factors induces specific changes in the kidney, known as nephrosclerosis. Arteriosclerotic fibro-intimal hyperplasia of afferent arterioles is the expected initial pathogenetic step of nephrosclerosis. This condition encompasses four primary aspects:

1. Arteriosclerosis, 2. Glomerulosclerosis, 3. Tubular Atrophy, and 4. Interstitial Fibrosis.

Age-related progressive nephrosclerosis reduces kidney mass, renal plasma flow (RPF), number of functioning nephrons, filtration membrane area, and glomerular capillary permeability. Therefore, the glomerular filtration rate (GFR) usually declines with age.^66–69^

### 1.4.1. Glomerular Hemodynamics in Aging

In the early stages of kidney damage, reduced vascular resistance in the afferent arterioles and increased responsiveness of the efferent arterioles to vasoconstrictive signals lead to elevated glomerular capillary hydrostatic pressure (GCHP).^66^ Despite the increased GCHP, the reactive reduction of glomerular capillary ultrafiltration coefficients prevents a corresponding increase in filtration efficiency and glomerular filtration rate (GFR). Initially, hyperfiltration may occur, but over time, the reduction in ultrafiltration coefficients and increased vasoconstriction of afferent arterioles also impair capillary filtration efficiency and GFR.^66,70^

#### 1.4.2. Age- and Sex-Related Decline of the Glomerular Filtration Rate (GFR)

The GFR is low at birth and increases to adult levels by the age of two years. It remains stable until around 40 and gradually decreases by about 5%–10% per decade.^68^ This decline is mainly due to the progressive loss of functioning nephrons. Women typically have lower GFR than men until around the sixth decade, when sex-disparities diminish and may reverse due to a faster decline in male GFR (1.20 vs. 0.96 ml/min per 1.73 m^2^ per year).^71^ This phenomenon is attributed to higher blood pressure and earlier arteriosclerotic cardiovascular disease in men.^68,71–76^ Hormonal factors additionally contribute to the gender difference in GFR decline as estrogens exert renoprotective effects by inhibiting the RAAS and facilitating advantages in methylation and antioxidation.^10,67,68,77,78^

By contrast, testosterone tends to increase potentially harmful RAAS activity, and androgen receptors mediate cellular aging by telomere shortening and induction of tubular cell apoptosis (Supplements 3 and 6).^10,77,78^ Consistently, experimental estrogen therapy and androgen deprivation could be demonstrated to mitigate the progression of chronic kidney disease (CKD).^66^

In advanced stages, the decline in GFR is accompanied by increased permeability of the glomerular basement membrane to macromolecules, facilitating the urinary excretion of significant amounts of albumin and other proteins.^66,79^

#### 1.4.3. Tubular System Changes

Aging also affects the tubular system, leading to tubulointerstitial fibrosis and impairing the secretion and reabsorption of various urinary components. The loop of Henle gradually loses control over salt reabsorption, and the decreased sensitivity of distal tubules and collecting ducts to aldosterone and arginine vasopressin (AVP) results in increased urinary dilution and further reductions in urinary creatinine (CRN).^23,80,81^

#### 1.4.4. Impact of Aging on Renal Vascular Function

As individuals age, renal vascular function progressively declines due to atherosclerosis and altered hormonal vasoregulation. Enhanced responses to vasoconstrictors and reduced reactions to vasodilators in various resistance vessels increase total renal vascular resistance and reduce renal plasma flow (RPF), especially in hypertension and albuminuria.^82^ These changes result from increased vasoconstrictive activities of AVP and the renin-angiotensin-aldosterone system (RAAS) besides decreased nitric oxide (NO) activity, amplifying renal vasoconstriction, sodium retention, matrix production, and mesangial fibrosis. ^66,68,82–86^

#### 1.4.5. Creatinine Clearance and Creatinine in Serum and Urine

The decline in GFR with age leads to decreased creatinine clearance and lower 24-hour urinary CRN. However, spot urine measurements of CRN are unreliable estimates for creatinine clearance due to their strong dependence on hydration status. Similarly, serum creatinine is not a sensitive marker of kidney function in older adults. This is because their reduced muscle mass results in a lower creatinine (CRN) load to be excreted by the kidneys, and the increased secretion of creatinine in the tubules prevents a rise in serum CRN levels in the early stages of chronic kidney disease (CKD), despite reduced glomerular filtration rates (GFR).^50,66,87^

### 1.5. Influence of Age on Renal Arsenic Excretion

Numerous studies have investigated how age affects the renal excretion of arsenic. These studies have focused on measuring total weight urinary arsenic (TWuAs), including arsenobetaine, and the sum of inorganic arsenic and its primary metabolites, MMA and DMA (total urinary arsenic, TuAs), excluding arsenobetaine. Additionally, they have examined the concentration of individual arsenic species in urine in absolute terms and relative to total weight arsenic.^34,39,55,88–90^ A significant challenge in interpreting these studies is the issue of adequate dilution adjustment. Older adults generally exhibit lower CRN, urinary osmolality, and specific gravity due to reduced renal urine concentration capacity and muscle mass.^91^ This can lead to inaccuracies, with uncorrected urine values being lower due to higher dilution, while classically corrected values may be inadequately high due to the smaller corrective denominators.^2,3^

In a Korean study, arsenic urine levels of 1,507 participants were standardized using a covariate-adjusted method that accounted for the influences of age, sex, and BMI on CRN.^1,34,44^ This advanced approach revealed significant positive associations between age and TWuAs or arsenobetaine in adults over 18. Positive trends for iAs, MMA, and DMA were observed in adults over 65 but did not reach statistical significance. Another study of 904 participants in Nevada, which included CRN as an independent covariate in a multiple linear regression model, found that age significantly negatively influenced iAs and %iAs while showing positive but non-significant trends for MMA, DMA, %DMA, and TuAs.^55^ Age was positively associated with methylation capacity, significantly affecting the primary (PMI = MMA/iAs) and marginally affecting the secondary methylation index (SMI = DMA/MMA). These findings align with previous studies showing increased methylation of urinary arsenic in adults, with lower %iAs and higher %DMA.^39,88^

The reduction in %iAs with age may also be influenced by declines in the proteohormone transmembrane Klotho (m-Kl) associated with aging or chronic kidney disease (CKD).^56,92–94^ As an essential cofactor of fibroblast growth factor 23 (FGF23), m-Kl assists the urinary excretion of iAs_V_ by inactivating the apical tubular sodium-phosphate cotransporters NaPi-IIa (and NaPi-IIc) mentioned above.^95–97^ Lower activity of m-Kl in CKD increases the reabsorption of iAs_V_ and its cellular conversion into iAs_III_ and subsequent methylation. Dehydration, common in older adults, exacerbates this effect by activating AVP and ATII, which further reduces the excretion of PO_4_^−^ and iAs_V_.^98,99^ Serum levels of m-Kl are higher in females throughout life and are positively correlated with estrogen, testosterone, and SHBG in men.^57,58^

Contrary results were observed in Taiwan’s hyperendemic arseniasis area,^42^, where older individuals exhibited increased %iAs and decreased %DMA, suggesting regional differences in age-dependent urinary arsenic composition due to diverse exposure sources or genetic differences in methylation efficiency.^40^ A study in Bangladesh involving children found lower primary (PMI) and higher secondary methylation indices (SMI) in children than adults, further supporting age-dependent differences in methylation efficiency and potentially explaining the absence of arsenic-induced skin lesions in children.^89^

In individuals over 90, there may be a reversal in age-related inorganic arsenic (iAs) reabsorption, inferred from observed drops in serum phosphate (PO_4_^−^) levels in this age group, which tend to increase from the third decade of life.^53^ Age-related declines in renal concentrating ability and calcitriol (vitamin D3) formation lead to higher renal losses and reduced intestinal absorption of calcium.^57,58^ This increases plasma parathyroid hormone (PTH), potentially facilitating renal hyperexcretion of PO_4_^−^ and iAs_v_ via downregulating NaPi-IIa transporters.^56,92–94^ While in younger seniors, these effects are overcompensated by the age-related decline in membrane-Klotho (m-KL), in the very old, these processes may prevail and lead to increased PO_4_^−^ and iAs_v_ excretion. However, the biotransformation and renal handling of arsenic in this age group remains to be studied in more detail.^53^

### 1.6. Influence of Kidney Function on Arsenic Excretion

Chronic kidney disease (CKD) has a well-established association with urinary arsenic levels.^4,59,62^ Several studies have shown significant negative correlations between the estimated glomerular filtration rate (eGFR) and total urinary arsenic (TuAs), including inorganic arsenic and its primary metabolites, MMA and DMA.^4,64,100^ Additionally, there have been positive associations between TuAs and serum cystatin C,^101^ albuminuria, or proteinuria.^62^ Moreover, there are positive correlations between TuAs and urinary markers for tubular damage, such as β2-microglobulin and N-acetyl-β-D-glucosaminidase (NAG),^4,100^ and in various populations, CKD mortality rates consistently increase with rising TuAs levels.^4,102,103^

Studies investigating the influence of renal function on arsenic excretion, akin to those on age (Section 1.4), indicate a decrease in the percentage of inorganic arsenic (%iAs) alongside concurrent elevations of the percentage of dimethylarsinic acid (%DMA) as glomerular filtration declines.^60,61^ Furthermore, there is evidence for impaired excretion of inorganic arsenic from blood to urine in dysfunctional kidneys, as the corresponding elevation of urinary %iAs is diminished with reduced GFR for a given increase in blood %iAs.^60^

These findings support the interpretation as ‘reverse causation,’ where reduced GFR is understood as a causal factor for impaired renal excretion of inorganic arsenic.^61^ Prolonged retention of inorganic arsenic in the blood may facilitate its further methylation, leading to eventual excretion as dimethylarsinic acid. Biochemical evidence supports this notion. Specifically, the downregulation of membrane-Klotho (m-KL) results in higher reabsorption and lower excretion of phosphate and inorganic arsenic in chronic kidney disease (CKD) (See Section 1.4 and Supplement 3).^104^ In CKD, increased renal angiotensin II (ATII), due to renin release in response to reduced renal plasma flow, inhibits m-KL. The pharmacological blockade of ATII consistently upregulates m-KL expression,^105^ thereby counteracting the inhibitory influence of m-KL depletion on renal inorganic arsenic excretion.^106^

In advanced chronic kidney disease, the effects of m-KL reduction on total urinary arsenic (TuAs) composition may be augmented by concurrent tubular dysfunction. This may result in further shifts from inorganic arsenic (%iAs) to dimethylarsinic acid (%DMA) due to disproportionate alterations in the net excretion of various total urinary arsenic constituents.^3^

However, the concept of reverse causation in the observed associations between glomerular filtration rate and urinary arsenic and its methylated forms is challenged. A higher percentage of dimethylarsinic acid (%DMA) and a lower percentage of inorganic arsenic (%iAs) in CKD could potentially induce kidney damage rather than result from it.^4^ This is because the oxidative methylation of inorganic arsenic to dimethylarsinic acid generates more toxic intermediates, such as MMAIII or DMAIII, which have been inconsistently found to negatively correlate with the estimated glomerular filtration rate (eGFR).^4,107–110^

This study utilized a comprehensive dataset from diverse populations to elucidate the intricate influence of sex- and age-related variability of the metabolism of arsenic and CRN, kidney function, and fluid balance on arsenic levels in both blood and urine across various dilution adjustment modes.

## 2. Methodology

### 2.1. Study Population

The study was based on anonymized data of n = 5,752 spot urine samples (Set 1) collected between January 2014 and August 2022 from the Institute for Medical Diagnostics Berlin-Potsdam, Germany (IMD), as used in previous research for V-PFRC formula generation.^3^ The data included information on sex, age, CRN, and total weight of urinary arsenic, which was part of multi-element metal screening panels in the urine of diverse populations in Germany, Switzerland, and Austria. These panels were conducted as routine preventive and diagnostic medical screening procedures, ensuring a comprehensive and generally demographically representative dataset. Apart from sex and age, no further health-related clinical information was available from the Institute for Medical Diagnostics.

A further dataset (Set 2) of 1,170 samples was investigated to compare urinary results with blood, where total weight arsenic was detected in parallel in EDTA whole blood and urine. The samples of Set 2 were obtained and analyzed by IMD between April 2014 and March 2024. The original 1,175 samples were trimmed for CRN < 3.5 g/L and age ≤ 85. Of the remaining 1,154 samples, 70.4% were part of the first dataset, while 29.6% were collected later. The key statistics of Set 1 and Set 2 are summarized in Tables 1 and 2. Histograms comparing data distributions of both sets are depicted in Supplements 8–13. It can be assumed that the samples represent the combined background exposure of inorganic and organic arsenic, as there was no specific suspicion of arsenic contamination in drinking water leading to the multi-element metal screening. The recommended WHO limit of 10 µg/L arsenic in drinking water is enforced and generally met in the sample collection areas.^111–114^ Any significant elevations of total weight arsenic levels in these regions are most commonly attributable to arsenobetaine, mainly from marine food, considering the quantitative predominance of this organic source.^49,115^

**Table 1:**
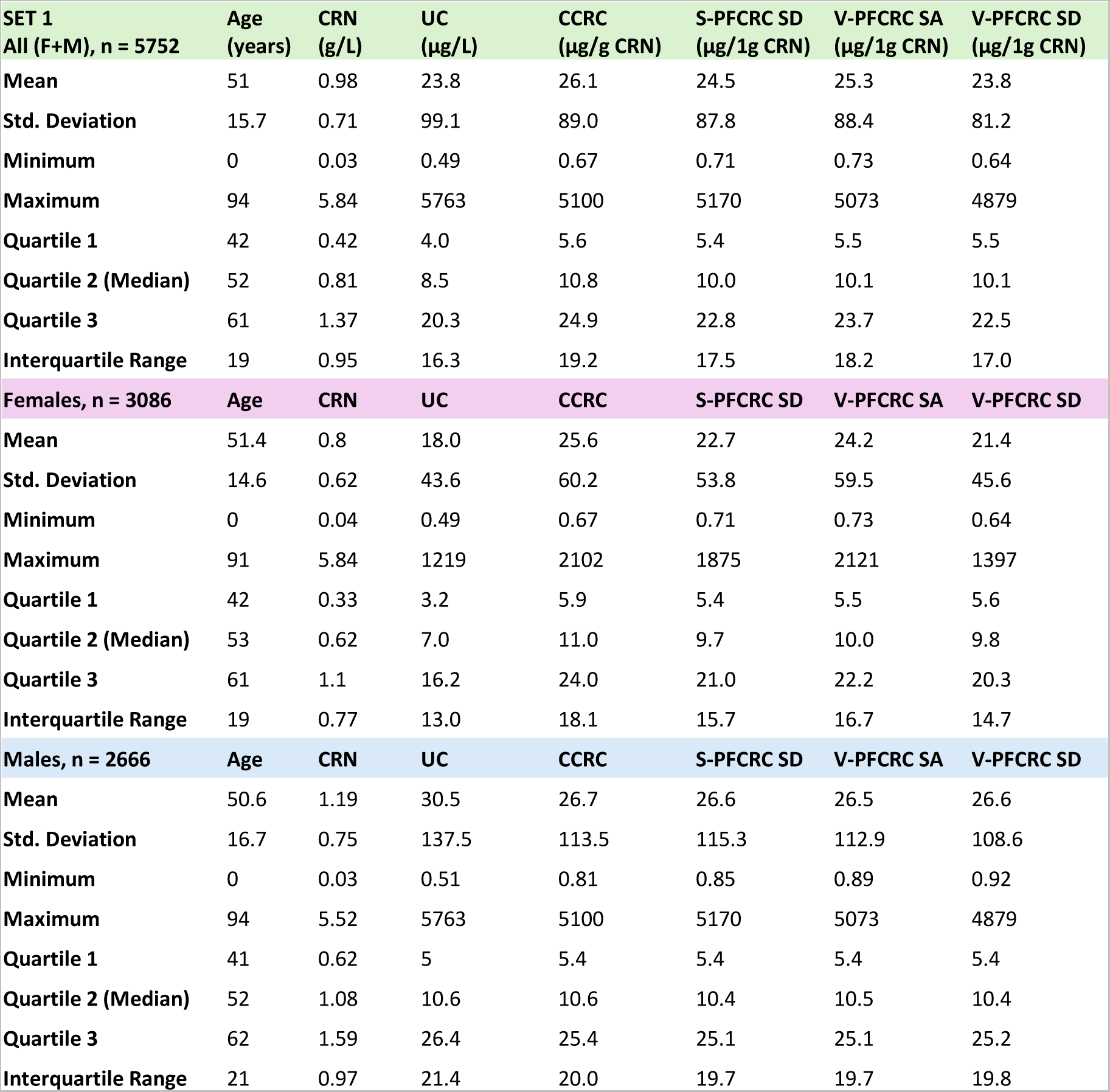
Basic statistics of Set 1. Sex-aggregated and -differentiated data are given for age, CRN, and urinary arsenic of the five result modes detailed in Table 3: Uncorrected (UC), Conventional CRN-correction (CCRC), Sex-differentiated simple power functional CRN correction (S-PFCRC SD), Sex-aggregated (V-PFCRC SA) and Sex-differentiated variable power functional CRN correction (V-PFCRC SD).

| SET 1<br>All (F+M), n = 5752 | Age<br>(years) | CRN<br>(g/L) | UC<br>($\mu\text{g/L}$) | CCRC<br>($\mu\text{g/g CRN}$) | S-PFCRC SD<br>($\mu\text{g/1g CRN}$) | V-PFCRC SA<br>($\mu\text{g/1g CRN}$) | V-PFCRC SD<br>($\mu\text{g/1g CRN}$) |
| --- | --- | --- | --- | --- | --- | --- | --- |
| Mean | 51 | 0.98 | 23.8 | 26.1 | 24.5 | 25.3 | 23.8 |
| Std. Deviation | 15.7 | 0.71 | 99.1 | 89.0 | 87.8 | 88.4 | 81.2 |
| Minimum | 0 | 0.03 | 0.49 | 0.67 | 0.71 | 0.73 | 0.64 |
| Maximum | 94 | 5.84 | 5763 | 5100 | 5170 | 5073 | 4879 |
| Quartile 1 | 42 | 0.42 | 4.0 | 5.6 | 5.4 | 5.5 | 5.5 |
| Quartile 2 (Median) | 52 | 0.81 | 8.5 | 10.8 | 10.0 | 10.1 | 10.1 |
| Quartile 3 | 61 | 1.37 | 20.3 | 24.9 | 22.8 | 23.7 | 22.5 |
| Interquartile Range | 19 | 0.95 | 16.3 | 19.2 | 17.5 | 18.2 | 17.0 |
| Females, n = 3086 | Age | CRN | UC | CCRC | S-PFCRC SD | V-PFCRC SA | V-PFCRC SD |
| Mean | 51.4 | 0.8 | 18.0 | 25.6 | 22.7 | 24.2 | 21.4 |
| Std. Deviation | 14.6 | 0.62 | 43.6 | 60.2 | 53.8 | 59.5 | 45.6 |
| Minimum | 0 | 0.04 | 0.49 | 0.67 | 0.71 | 0.73 | 0.64 |
| Maximum | 91 | 5.84 | 1219 | 2102 | 1875 | 2121 | 1397 |
| Quartile 1 | 42 | 0.33 | 3.2 | 5.9 | 5.4 | 5.5 | 5.6 |
| Quartile 2 (Median) | 53 | 0.62 | 7.0 | 11.0 | 9.7 | 10.0 | 9.8 |
| Quartile 3 | 61 | 1.1 | 16.2 | 24.0 | 21.0 | 22.2 | 20.3 |
| Interquartile Range | 19 | 0.77 | 13.0 | 18.1 | 15.7 | 16.7 | 14.7 |
| Males, n = 2666 | Age | CRN | UC | CCRC | S-PFCRC SD | V-PFCRC SA | V-PFCRC SD |
| Mean | 50.6 | 1.19 | 30.5 | 26.7 | 26.6 | 26.5 | 26.6 |
| Std. Deviation | 16.7 | 0.75 | 137.5 | 113.5 | 115.3 | 112.9 | 108.6 |
| Minimum | 0 | 0.03 | 0.51 | 0.81 | 0.85 | 0.89 | 0.92 |
| Maximum | 94 | 5.52 | 5763 | 5100 | 5170 | 5073 | 4879 |
| Quartile 1 | 41 | 0.62 | 5 | 5.4 | 5.4 | 5.4 | 5.4 |
| Quartile 2 (Median) | 52 | 1.08 | 10.6 | 10.6 | 10.4 | 10.5 | 10.4 |
| Quartile 3 | 62 | 1.59 | 26.4 | 25.4 | 25.1 | 25.1 | 25.2 |
| Interquartile Range | 21 | 0.97 | 21.4 | 20.0 | 19.7 | 19.7 | 19.8 |

**Table 2:** Basic statistics of the entire Set 2. Sex-aggregated and -differentiated data are given for age, CRN, and arsenic in EDTA blood and urine in the five result modes detailed in Table 3.

| SET 2<br>All (F+M), n = 1170 | Age<br>(years) | CRN<br>(g/L) | UC<br>(µg/L) | CCRC<br>(µg/g CRN) | S-PFCRC SD<br>(µg/1g CRN) | V-PFCRC SA<br>(µg/1g CRN) | V-PFCRC SD<br>(µg/1g CRN) | EDTA Blood<br>(µg/L) |
| --- | --- | --- | --- | --- | --- | --- | --- | --- |
| Mean | 51.6 | 1.12 | 33.2 | 29.8 | 29.1 | 29.2 | 28.5 | 1.7 |
| Std. Deviation | 14.21 | 0.77 | 112.0 | 79.8 | 81.4 | 79.1 | 74.9 | 3 |
| Minimum | 5.6 | 0.04 | 0.4 | 0.8 | 0.8 | 0.9 | 0.9 | 0.08 |
| Maximum | 87.9 | 5.84 | 3176 | 2146 | 2241 | 2133 | 1917 | 44 |
| Quartile 1 | 42.3 | 0.54 | 4.3 | 5.3 | 5.2 | 5.2 | 5.3 | 0.4 |
| Quartile 2 (Median) | 52.2 | 0.96 | 10.2 | 10.8 | 10.6 | 10.6 | 10.5 | 0.8 |
| Quartile 3 | 60.4 | 1.53 | 26.4 | 29.5 | 27.9 | 28.8 | 26.9 | 1.8 |
| Interquartile Range | 18.1 | 0.99 | 22.1 | 24.2 | 22.7 | 23.6 | 21.6 | 1.4 |
| Females, n = 596 | Age | CRN | UC | CCRC | S-PFCRC SD | V-PFCRC SA | V-PFCRC SD | EDTA Blood |
| Mean | 51.5 | 0.95 | 22.7 | 26.3 | 24.5 | 25.0 | 24.0 | 1.4 |
| Std. Deviation | 14.2 | 0.72 | 52.3 | 51.6 | 49.9 | 50.3 | 49.4 | 2.9 |
| Minimum | 5.7 | 0.04 | 0.4 | 1.1 | 1.2 | 1.1 | 1.0 | 0.10 |
| Maximum | 84.8 | 5.84 | 778 | 778 | 778 | 778 | 778 | 44 |
| Quartile 1 | 42.4 | 0.4 | 3.3 | 5.2 | 4.8 | 4.9 | 5.0 | 0.3 |
| Quartile 2 (Median) | 51.8 | 0.78 | 7.7 | 9.6 | 9.1 | 9.4 | 9.1 | 0.7 |
| Quartile 3 | 59.7 | 1.3 | 19.3 | 26.4 | 22.7 | 23.1 | 21.9 | 1.5 |
| Interquartile Range | 17.3 | 0.9 | 16.0 | 21.2 | 18.0 | 18.2 | 16.9 | 1.2 |
| Males, n = 574 | Age | CRN | UC | CCRC | S-PFCRC SD | V-PFCRC SA | V-PFCRC SD | EDTA Blood |
| Mean | 51.8 | 1.29 | 44.2 | 33.4 | 33.8 | 33.6 | 33.1 | 2.0 |
| Std. Deviation | 14.3 | 0.78 | 150.0 | 101.0 | 104.4 | 100.6 | 94.1 | 3.1 |
| Minimum | 5.6 | 0.12 | 0.6 | 0.8 | 0.8 | 0.9 | 0.9 | 0.08 |
| Maximum | 87.9 | 4.48 | 3176 | 2146 | 2241 | 2132 | 1917 | 29 |
| Quartile 1 | 42.2 | 0.67 | 5.5 | 5.6 | 5.7 | 5.8 | 5.7 | 0.5 |
| Quartile 2 (Median) | 53.1 | 1.19 | 13.0 | 12.1 | 11.9 | 11.9 | 11.9 | 1.0 |
| Quartile 3 | 60.8 | 1.73 | 37.6 | 33.5 | 32.9 | 32.9 | 33.3 | 2.3 |
| Interquartile Range | 18.6 | 1.06 | 32.1 | 27.9 | 27.2 | 27.1 | 27.6 | 1.8 |

### 2.2. Treatment of Arsenic Values Below the Detection Limit

The 116 female and 32 male samples displaying uncorrected urinary arsenic levels below the ICP-MS detection limit of 1 µg/L were assumed to range between 0.5 and 0.99 µg/L due to prevalent arsenic exposure rendering values 0.5µg/L improbable. Values below the detection limit were estimated considering the apparent strong positive association of uncorrected arsenic and CRN using the formulas:

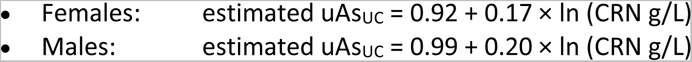

This method distributed samples within the 0.5-0.99 µg/L range, averaging 0.62 µg/L for females and 0.72 µg/L for males. This approach provided more plausible distributions than omitting values or using the usual fixed substitutions like half the detection limit (0.5 µg/L) or the limit divided by √2 (0.71 µg/L). For the 69 blood samples below the 0.2 µg/L detection limit, values were estimated based on the strong association of CRN-corrected urinary with blood arsenic using the unisex formula:

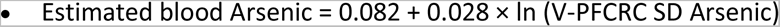

This approach distributed blood values between 0.08 and 0.19 µg/L, averaging 0.12 µg/L (60% of the detection limit).

### 2.3. Sample Collection, Preservation, Transportation, and Storage Procedures

#### 2.3.1. Urine

Urine samples were collected in neutral monovettes (Sarstedt) and shipped to the laboratory at room temperature within 24 hours for immediate CRN measurement. They were then stored at 4°C for 1-4 days before ICP-MS measurement.

#### 2.3.2. Blood

Venous whole blood was collected in tubes with EDTA for anticoagulation (Sarstedt) and transported to the laboratory overnight or by surface mail. The stability of elements in these specimens was validated over 14 days at room temperature.

### 2.4. Sample Preparation and Analysis Procedures

#### 2.4.1. Inductively Coupled Plasma-Mass Spectrometry (ICP-MS) Measurements of Arsenic

The total weight of arsenic in anticoagulated whole blood and urine was determined using ICP-MS. Arsenic species were not analyzed. For ICP-MS analysis, urine samples were diluted 1:10 in 1% HNO3 (Suprapur, Supelco), and serum and anticoagulated whole blood samples were diluted 1:20 in 0.1% NH3 (Suprapur, Supelco) and 0.02% Lutrol F88 (AppliChem). Metals were analyzed in collision/reaction cell mode using the ICapQ ICP-MS system (Thermo Fisher), with external and internal standard calibration (Elemental Scientific). Results are the means of three measurements.

#### 2.4.2. Creatinine Measurement

Creatinine analysis was performed enzymatically using the Alinity assay (Abbott Laboratories).

### 2.5. Normalization of Arsenic to Urinary Concentrations of 1 g/L CRN

The standardization of CRN-corrected urine measurements to 1g/L CRN and the process of V-PFCRC formula generation are explained in detail in previous work.^3^ Two general forms of power-functional correction can be distinguished:

1. The simple power functional CRN-correction (S-PFCRC) employing a fixed exponent B in the corrective formula:

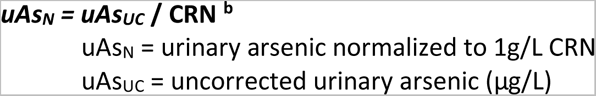 Uncorrected and conventionally CRN-corrected (CCRC) results can be seen as particular cases of S-PFCRC, with the uncorrected results taking the exponent b as 0 and the CCRC as 1.
2. The variable power functional correction (V-PFCRC) using two analyte-specific coefficients, c, and d, allowing the calculation of a variable exponent b depending on the concentration of the analyte and CRN:

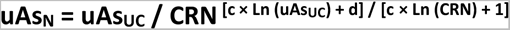

Both the simple and variable power functional correction formulae were adopted from the previous study.^3^ The simple formula was tested in a sex-differentiated mode, using separate exponents b for males and females. The variable formula was tested in sex-differentiated and sex-aggregated modes, using identical c and d values in both sexes. For all power-functional modes tested in this paper, the asymmetrical distribution of CRN values was compensated as described in the previous study, as asymmetry-compensation improved the accuracy of V-PFCRC.^3^ Table 3 summarizes the exponents b (for S-PFCRC) and the coefficients c and d (for V-PFCRC) used.

**Table 3:** Exponents and coefficients used in corrective formulas.^3^.

| Result Mode | Sex-differentiate or aggregate SD / SA | Exponent b (S-PFCRC) |  | Coefficient c (V-PFCRC) |  | Coefficient d (V-PFCRC) |  |
| --- | --- | --- | --- | --- | --- | --- | --- |
|  |  | Female | Male | Female | Male | Female | Male |
| Uncorrected | – | 0 |  | – |  |  |  |
| CCRC | – | 1 |  | – |  |  |  |
| V-PFCRC | SD | – |  | – 0.11 | 0.79 | 1.05 | 0.69 |
|  | SA | – |  | 0.03 |  | 0.78 |  |
| S-PFCRC | SD | 0.79 | 0.89 | – |  |  |  |

### 2.6. Statistical Analysis

Data were analyzed using Excel and Datatab.net software. Results were expressed as mean ± standard error, medians, and quartile limits (Q1 and Q3). The normality of data distribution was evaluated using the Kolmogorov–Smirnov and Q-Q plots. For data with normal or near-normal distribution, 2-sided Student’s t-tests and Pearson correlation analysis were used. Mann-Whitney U-tests were applied to test the significance of non-normally distributed data, and Spearman rank correlation was used for analysis. A p-value of <0.05 was considered statistically significant.

#### 2.6.1 Analysis of Age- and Sex-Effects on Arsenic in Blood and Urine

For the age and gender-stratified analysis, urine results from 5,752 samples in set 1 and urine and blood results from 1,154 samples in set 2 were analyzed for their association with age and residual dependence on CRN using Spearman correlation (r and p-value). For urine findings from set 2, Spearman correlations were also determined for corresponding EDTA blood arsenic. The Spearman ranking method was chosen due to the non-normal distribution of all linear and log-transformed data sets (CRN, blood, and urine values).

To assess the age effect independently of the asymmetrical age distribution, samples in set 1 were divided into nine ten-year age bands. Each band was percentile ranked separately for both sexes, excluding the top and bottom 5% of values to limit outlier distortion. The remaining 5,166 samples (2,773 female and 2,393 male) were analyzed age stratified. Basic statistics (number, mean, standard deviation, standard error, minimum, maximum, and quartiles 1-3) for age, creatinine, and all urinary outcome modes, as summarized in Table 3, were calculated separately for each ten-year age band. Results are provided in the Supplementary Excel file, and age kinetics of all outcome modes are visualized in Figure 1 using mean values and standard errors. The significance of sex differences was determined using a two-sided Student’s t-test on near-normally distributed logarithmized data sets.

**Figure 1:**
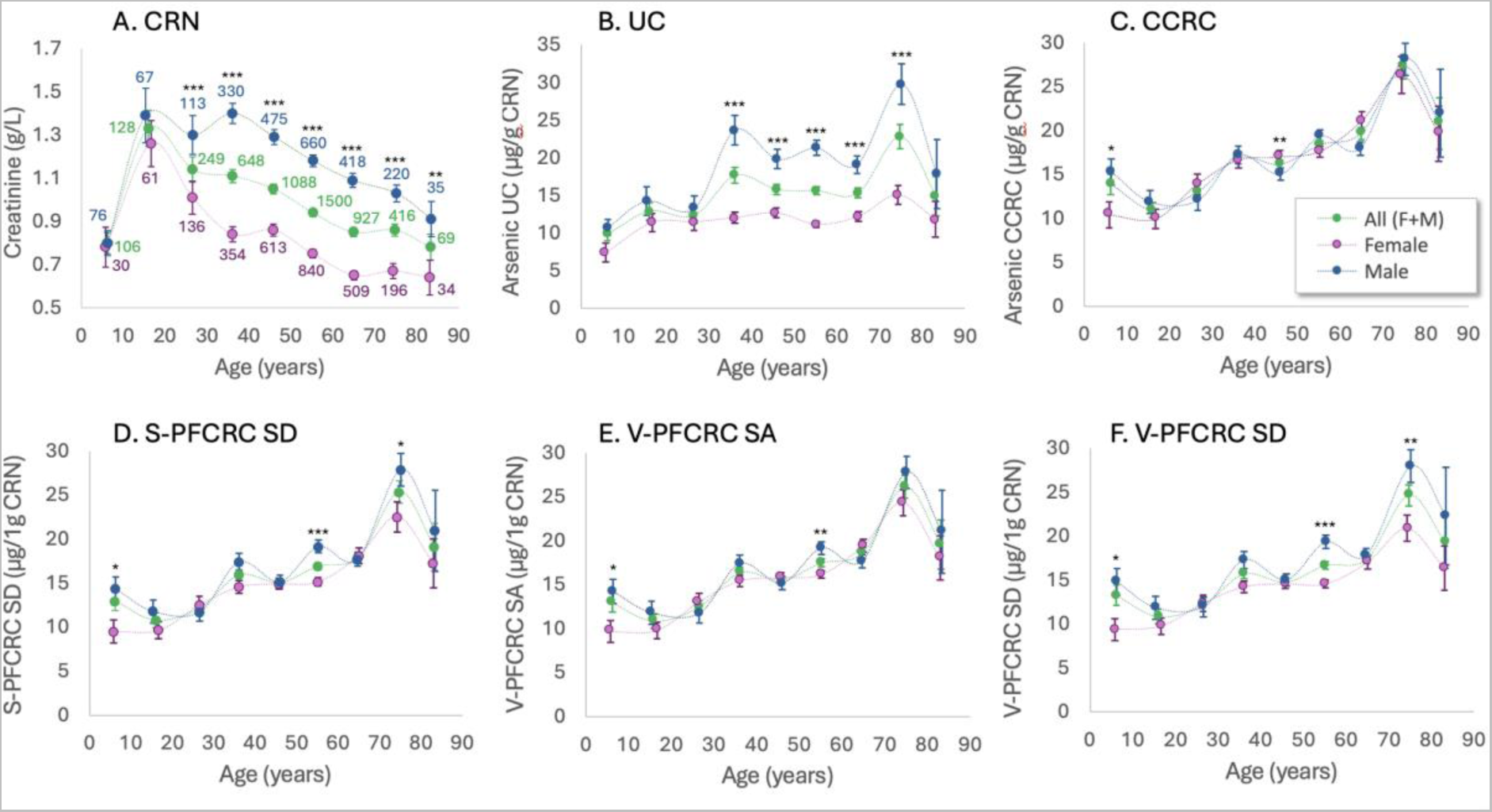
Sex-Differentiated Influence of Age on CRN and Urinary Arsenic across five result modes in Set 1. Means ± standard errors are plotted for nine 10-year age bands. Outliers above the 95% and below the 5% percentile of each age band were not analyzed. Post-outlier exclusion sample sizes are listed in Graph A. Urinary result modes are detailed in Section 2.5. Group statistics are provided in the Supplementary Excel file. Significance levels of sex differences (2-sided t-test) : * 0.05 ≥ p > 0.025, ** 0.025 ≥ p > 0.01, *** p < 0.001.

Similar evaluation procedures were performed for Set 2. Due to a substantially lower sample number than Set 1 and insufficient samples for the youngest and oldest patients, samples from patients aged 21 to 80 were divided into four 15-year bands, each trimmed to the 5-95 percentile. Urine and blood findings (uncorrected, CCRC, and the most effective sex-differentiated V-PFCRC SD mode) were evaluated analog to Set 1.

To illustrate the effects of age on arsenic concentrations in blood and urine result modes (UC, CCRC, and V-PFCRC-SD), the percentage deviations of quartiles 1-3 for the three older age groups from the values of the youngest age group in Set 2 were calculated. The results were presented in sex-differentiated and -aggregated bar charts.

#### 2.6.2. Determination of Age and Sex-specific V-PFCRC-Formulae

To investigate the impact of age and gender on the formulas, the sample population was divided into age groups: 18-45, 46-60, and 61-85. Vital statistical data for these groups are summarized in the Supplementary Excel file. For each age group and both sexes, the relationship between the exponents (b) and the V-PFCRC-normalized arsenic value (uAsN) was examined separately to determine analyte-specific coefficients (c and d), as described in the initial study.^3^ To minimize the impact of extreme values, the top 1% of CRN-stratified TWuAs values were excluded from determining V-PFCRC formulae. Due to the comparatively small number of samples, CRN asymmetry compensation was not performed. Instead, the concentration range was restricted to CRN values less than 3 g/L. This restriction was based on previous findings, resulting in significant alignment of the uncorrected and the asymmetry-compensated curves.^3^ The six age-sex-specific log-linear regression curves obtained were plotted and compared within and between both sexes, as shown in Figure 6.

#### 2.6.3. Validation of Standardization Efficacy in Separate Age Groups

The V-PFCRC and S-PFCRC formulas used in this study were initially developed for a broad age group ranging from 14 to 82 years. To evaluate and compare their age-specific standardization efficacy, study Set 1 was divided into seven age bands: 0-10, 11-20, 21-35, 36-50, 51-65, 66-80, and over 80. A power functional regression analysis of the type Arsenic = a x CRN^b^ was performed for each age band to determine the relationship between creatinine (CRN) and urinary arsenic. The slopes (b) and R² values of the resulting regression curves were used to measure the residual dependence on CRN for the five outcome modes detailed in Table 3. Lower values in both metrics indicated higher CRN independence, suggesting better equalization efficiency. These regression results were separately compared using bar charts for each age group and aggregated for the entire Set 1. Additional age-aggregated analyses were conducted for both sexes separately.

Similarly, the same five outcome modes from Set 2 were analyzed to evaluate the agreement between blood and urine results. The coefficients of determination (R²) of the regression curves of the power function between the EDTA blood values and the respective urine results were calculated for the five age groups: 0-20, 21-35, 36-50, 51-65, and 66-80. Additionally, the mean R² values of these five groups were calculated, and the total population of Set 2, aged 5-85, was analyzed. A high R² in this context indicates a strong correlation between the arsenic levels in blood and urine, demonstrating effective dilution adjustment of the urine results. A 66-year-old patient with implausibly low urine values and very high blood arsenic concentrations was excluded from this analysis. The statistics of Sets 1 and 2 age groups are summarized in the Supplementary Excel file.

## 3. Results

### 3.1. Effects of Age and Sex on CRN and Urinary Arsenic

Figures 1a-f illustrate how age and sex influence CRN and urinary arsenic levels across five result modes in Set 1. Creatinine levels increase steadily in children and adolescents due to growing lean muscle mass, peak in early adulthood, and gradually diminish with age. This age decline is more pronounced in males (r = −0.14) than in females (r = −0.1), starting later in males between the second and third decades. For females, the decrease begins shortly after puberty, influenced by generally lower lean muscle mass and the sex hormonal influence on fluid metabolism outlined in Section 1.2.

In contrast, arsenic levels show an age-related rise for both sexes across all urine result modes, indicating actual age-related exposure increments beyond the effects of declining urinary CRN. The correlation with age for CRN-corrected arsenic levels is weak but significant: overall r = 0.11–0.12, females r = 0.09–0.11, and males r = 0.13–0.14. Linear regression analysis indicates an average annual increase in V-PFCRC arsenic from puberty to the mid-seventies of 0.23 µg/g CRN for males and 0.16 µg/g CRN for females.

In CCRC urine, the lower CRN levels in females computationally amplify the apparent increase in arsenic more than in males, effectively narrowing the gender gap observed in uncorrected urinary arsenic (see Figures 1b and c). The effect is less pronounced in the power-functional CRN corrections, particularly in the sex-differentiated V-PFCRC SD, which retains a significant male bias in urinary arsenic. This residual gender difference in simple and variable PFCRC reflects a more accurate representation of actual exposure, as this gap is also present in the EDTA blood of Set 2 (Figure 2b). Consequently, the evidence supports the use of V-PFCRC SD and, to a lesser extent, the other two power-functional correction modes as providing more accurate estimations of exposure compared to CCRC.

**Figure 2:**
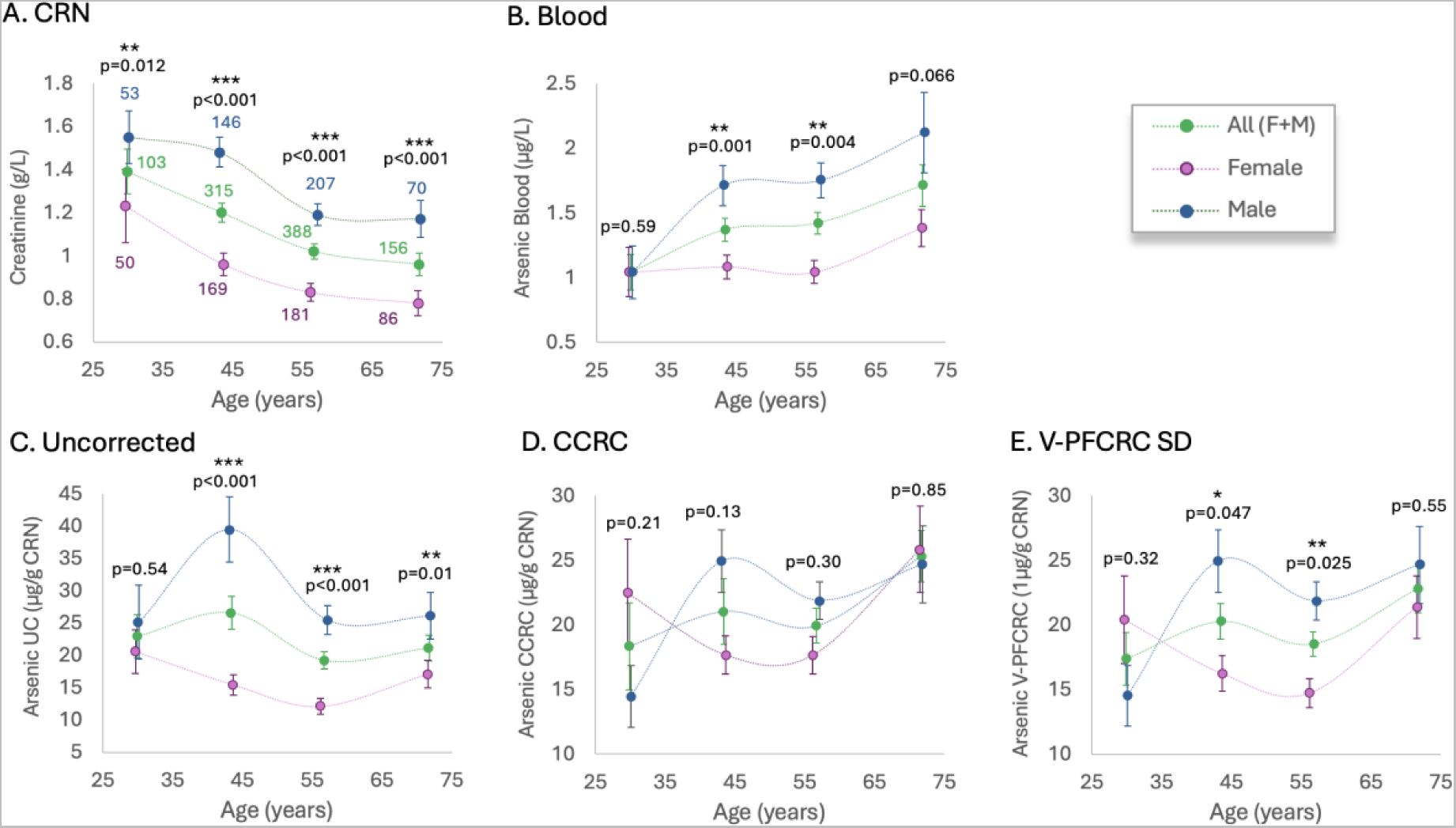
Sex-Differentiated Influence of Age on CRN, Urine, and Blood Arsenic in Set 2. Analogous representation to Figure 1 for Data Set 2, including arsenic values in EDTA blood determined in parallel with urine of three result modes in four 15-year-age bands (21–35, 36–50, 51–65, and 66–80 years). Means ± standard errors for the remaining 486 female and 476 male samples are plotted. Significance level of sex-difference (2-sided t-tests): * 0.05 > p > 0.025, ** 0.025 ≥ p > 0.01, *** p < 0.001.

Males consistently exhibit higher creatinine levels across all adult age groups, a disparity not seen in younger children and adolescents. Corresponding sex differences in uncorrected arsenic (uAs_UC_) lag those of CRN and are partly confounded by the generally higher urine concentration of males explained in Section 1.2. The sex differences in uAs_UC_ emerge in young adulthood and peak in the mid-forties due to steeper increases in males generating significantly higher uAs_UC_ across all adult age bands except the third decade. Consistently low but significant positive correlations between uAs_UC_ and age are observed in the combined and male data but not in females (overall r = 0.05, males r = 0.08).

Notably, both sexes experience steep increases in arsenic levels between the seventh and eighth decades. This late-life urinary arsenic flush, in the absence of corresponding drops in CRN, suggests actual overexposure to arsenic in this age group. This finding is supported by the highest arsenic levels in EDTA blood in both sexes at older ages, likely due to the redistribution of stored arsenic from internal sources.

### 3.2. Comparison of the Effects of Age and Sex on Urinary and Blood Arsenic

The relationship between age and arsenic levels in blood and urine was analyzed using a smaller dataset (Set 2), which included limited samples from patients under 20 and over 80.m Therefore, the age range of 20 to 80 was divided into four 15-year intervals. In line with previous research,^116,117^ average arsenic levels in blood were substantially lower than in urine, with V-PFCRC urine concentrations being 17.5 times higher in women and 16.6 times higher in men compared to EDTA whole blood. For women, blood arsenic levels remained stable across the first three age groups, increasing significantly only in the oldest group. Conversely, men showed a more continuous increase in blood arsenic levels from the youngest to the oldest age group, leading to significant male-dominated gender differences in the three older groups.

Both uncorrected and V-PFCRC results in Set 2 widely reflect the gender asymmetry and age dependence seen in Set 1. At the same time, CCRC adjustment consistently reduced this gender disparity to insignificance in both sets. Unlike the age-increasing kinetic of Set 1, women in Set 2 exhibited a trough-shaped trend in urinary arsenic levels, with an initial decrease in the younger age bands followed by an increase in the sixth decade of life. Men displayed an initial increase between the first and second groups, a decrease between the second and third groups, and a subsequent rise towards the oldest group. These discontinuous trends resulted in positive correlations between age and urinary arsenic, though less pronounced than those in Set 1 or those between age and blood arsenic, as detailed in Tables 4 and 5.

**Table 4:** Correlations between Age, CRN, and all urinary arsenic result modes of Set 1. Spearman coefficients r and significances p are given for non-normally distributed data.

| Set 1 / 21-94 Years |  |  |  |  |  |  |  |  |
| --- | --- | --- | --- | --- | --- | --- | --- | --- |
| All (F+M) | n = 5488 | Age | CRN | UC | CCRC | S-PFCRC SD | V-PFCRC SA | V-PFCRC SD |
| Age | Correlation |  | -0.11 | 0.04 | 0.11 | 0.11 | 0.1 | 0.1 |
|  | p |  | <.001 | 0.004 | <.001 | <.001 | <.001 | <.001 |
| CRN (g/L) | Correlation | -0.11 |  | 0.53 | -0.12 | 0.01 | -0.01 | 0.01 |
|  | p | <.001 |  | <.001 | <.001 | 0.591 | 0.625 | 0.522 |
| Female | n = 2983 | Age | CRN | UC | CCRC | S-PFCRC SD | V-PFCRC SA | V-PFCRC SD |
| Age | Correlation |  | -0.1 | 0.02 | 0.09 | 0.08 | 0.08 | 0.08 |
|  | p |  | <.001 | 0.405 | <.001 | <.001 | <.001 | <.001 |
| CRN (g/L) | Correlation | -0.1 |  | 0.56 | -0.13 | 0.03 | -0.01 | 0.03 |
|  | p | <.001 |  | <.001 | <.001 | 0.08 | 0.713 | 0.154 |
| Male | n = 2505 | Age | CRN | UC | CCRC | S-PFCRC SD | V-PFCRC SA | V-PFCRC SD |
| Age | Correlation |  | -0.14 | 0.06 | 0.14 | 0.13 | 0.13 | 0.13 |
|  | p |  | <.001 | 0.002 | <.001 | <.001 | <.001 | <.001 |
| CRN (g/L) | Correlation | -0.14 |  | 0.46 | -0.11 | -0.04 | -0.01 | -0.03 |
|  | p | <.001 |  | <.001 | <.001 | 0.065 | 0.516 | 0.185 |

**Table 5:** Spearman Correlations and significances between age and arsenic in all urinary arsenic result modes and blood of Set 2.

| Set 2 / 21-88 Years |  |  |  |  |  |  |  |  |  |
| --- | --- | --- | --- | --- | --- | --- | --- | --- | --- |
| All (F+M) | n = 1144 | Age | CRN | UC | CCRC | S-PFCRC SD | V-PFCRC SA | V-PFCRC SD | Blood |
| Age | Correlation |  | -0.15 | -0.01 | 0.08 | 0.06 | 0.07 | 0.06 | 0.1 |
|  | p |  | <.001 | 0.724 | 0.009 | 0.033 | 0.027 | 0.03 | <.001 |
| CRN (g/L) | Correlation | -0.15 |  | 0.5 | -0.09 | 0.03 | 0.01 | 0.02 | -0.03 |
|  | p | <.001 |  | <.001 | 0.003 | 0.374 | 0.706 | 0.449 | 0.373 |
| Blood | Correlation | 0.1 | -0.03 | 0.67 | 0.81 | 0.81 | 0.81 | 0.81 |  |
|  | p | <.001 | 0.373 | <.001 | <.001 | <.001 | <.001 | <.001 |  |
| Female | n = 582 | Age | CRN | UC | CCRC | S-PFCRC SD | V-PFCRC SA | V-PFCRC SD | Blood |
| Age | Correlation |  | -0.13 | -0.02 | 0.08 | 0.06 | 0.06 | 0.06 | 0.1 |
|  | p |  | 0.002 | 0.662 | 0.064 | 0.156 | 0.125 | 0.152 | 0.013 |
| CRN (g/L) | Correlation | -0.13 |  | 0.52 | -0.16 | 0 | -0.05 | -0.02 | -0.12 |
|  | p | 0.002 |  | <.001 | <.001 | 0.954 | 0.271 | 0.65 | 0.003 |
| Blood | Correlation | 0.1 | -0.12 | 0.58 | 0.79 | 0.77 | 0.78 | 0.78 |  |
|  | p | 0.013 | 0.003 | <.001 | <.001 | <.001 | <.001 | <.001 |  |
| Male | n = 562 | Age | CRN | UC | CCRC | S-PFCRC SD | V-PFCRC SA | V-PFCRC SD | Blood |
| Age | Correlation |  | -0.19 | -0.02 | 0.07 | 0.06 | 0.06 | 0.06 | 0.1 |
|  | p |  | <.001 | 0.71 | 0.093 | 0.147 | 0.155 | 0.128 | 0.022 |
| CRN (g/L) | Correlation | -0.19 |  | 0.44 | -0.05 | 0.01 | 0.03 | 0.02 | -0.02 |
|  | p | <.001 |  | <.001 | 0.273 | 0.764 | 0.463 | 0.701 | 0.598 |
| Blood | Correlation | 0.1 | -0.02 | 0.73 | 0.84 | 0.84 | 0.83 | 0.83 |  |
|  | p | 0.022 | 0.598 | <.001 | <.001 | <.001 | <.001 | <.001 |  |

Figure 2 shows that the V-PFCRC age-sex dynamics align more closely with blood arsenic levels than the CCRC.

### 3.3. Differential Effects of Aging on Arsenic in Blood and Urine

As shown in the previous section, mean arsenic concentrations in blood and urine increase with age in both sexes, though the kinetics vary depending on sex, test medium, and result mode. Figure 3 illustrates the relative age changes of three older groups compared to the youngest group (21-35 years). Differentiation according to quartile limits allows for investigating the influence of arsenic exposure on age-related changes in arsenic levels. In blood, the oldest age group shows the most significant increase in arsenic concentrations for both sexes and across all quartiles compared to the younger reference group. This increase in older age is higher than the blood arsenic increases observed in the two middle-aged groups, and proportionally stronger than the corresponding increase in all three urine modes of the oldest group.

**Figure 3:**
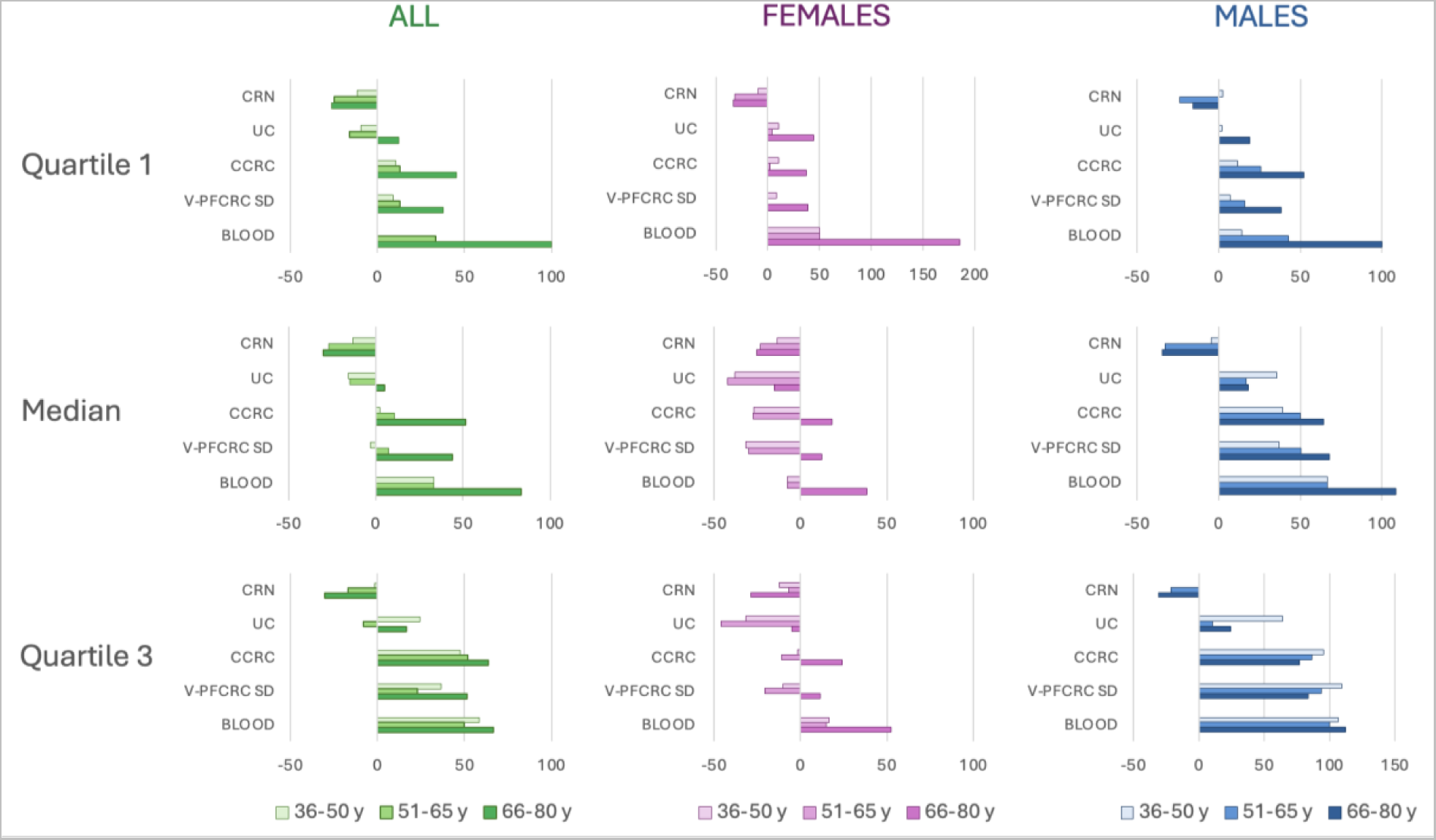
Comparison of Age Effects on Arsenic Concentrations in Blood and Urine. Percentage deviations of medians and quartile (Q1 and Q3) limits of the three older age groups of Set 2 (36-50,51-65, and 66-80) from the youngest age group (21-35 years) are plotted for blood and urine in UC, CCRC, and V-PFCRC mode.

Age-related changes of uncorrected urine in both sexes show the most significant discrepancies from blood. Arsenic in CCRC and V-PFCRC mode exhibit comparatively stronger agreement with blood values across all quartiles except for the female Q1. The alignment between urine and blood age kinetics, being generally more robust in men, improves in both sexes from quartile 1 through the median to quartile 3. Women show consistent age-related increases or parity in quartile 1 across all groups. In contrast, the median and Q3 values of the two middle-aged groups even show a reduction in arsenic levels compared to the youngest group. This suggests that gender differences in arsenic metabolism and excretion may be more pronounced at higher arsenic exposures. These findings highlight the complex interplay between age, sex, and arsenic metabolism, underscoring the importance of considering these factors in assessing arsenic exposure.

### 3.4. Age-Specific Sex Ratios of Quartile Limits 1-3

To investigate the extent to which the sex ratios in the individual urine result modes correspond to the blood ratios, the sex ratios of arsenic quartile limits 1-3 for blood and the three urine modes of Set 2 (UC, CCRC, and V-PFCRC SD) were calculated and compared.

Figure 4 illustrates the higher arsenic levels in males across most age groups and parameters represented by Female/Male ratios predominantly taking values < 1. Significant exceptions are observed in the group of 21-35-year-olds, where female CCRC and V-PFCRC urine values for medians and quartile 3 are significantly higher than those of males, and sex disparities in quartile 1 are broadly neutralized. The robustness of this inversion of sex ratios is supported by its consistent presence in the 20 to 30-year-old group (but not in the 31-40-year-olds) of the more extensive Set 1, as illustrated in Figure 1 and the detailed group statistics provided in the Supplementary Excel File.

**Figure 4:**
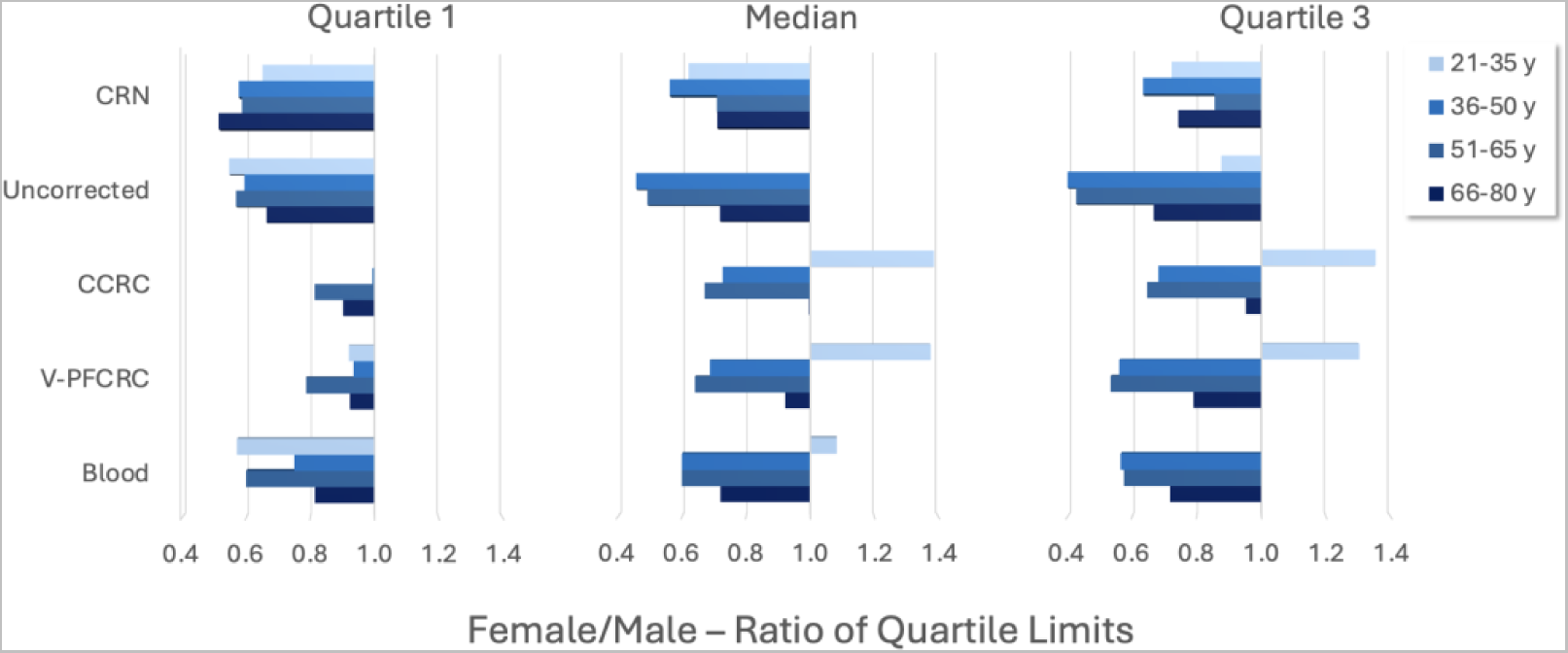
Sex-Ratios of Quartile Limits for CRN, Blood- and Urine Arsenic.

These urine findings correspond to the narrow female dominance in the median, gender neutrality in quartile 3, and converse male dominance in quartile 1 of blood arsenic in this age group. This suggests a gender paradoxical handling of arsenic limited to younger age groups and more pronounced in medium to higher arsenic exposures. Since hormonal differences are most apparent in this age group, a potential connection with endocrine factors is evident.

Figure 5 illustrates the percentage deviations of urinary from blood sex ratios of arsenic in Quartiles 1-3. The most minor deviations between blood and urine are found in uncorrected urine, while the most considerable deviations occur in CCRC. Compared to CCRC, the analysis indicates that V-PFCRC provides a better match for all quartiles and age groups, except for the 1st quartile in the 66-80 age group, where CCRC has a slight advantage. The most pronounced deviations are seen in the youngest group’s 1st quartile of CCRC and V-PFCRC, where urinary female/male ratios are significantly higher than those in blood. This suggests that young women may excrete arsenic more effectively in urine than men, possibly due to their greater efficiency of arsenic methylation related to estrogen.

**Figure 5:**
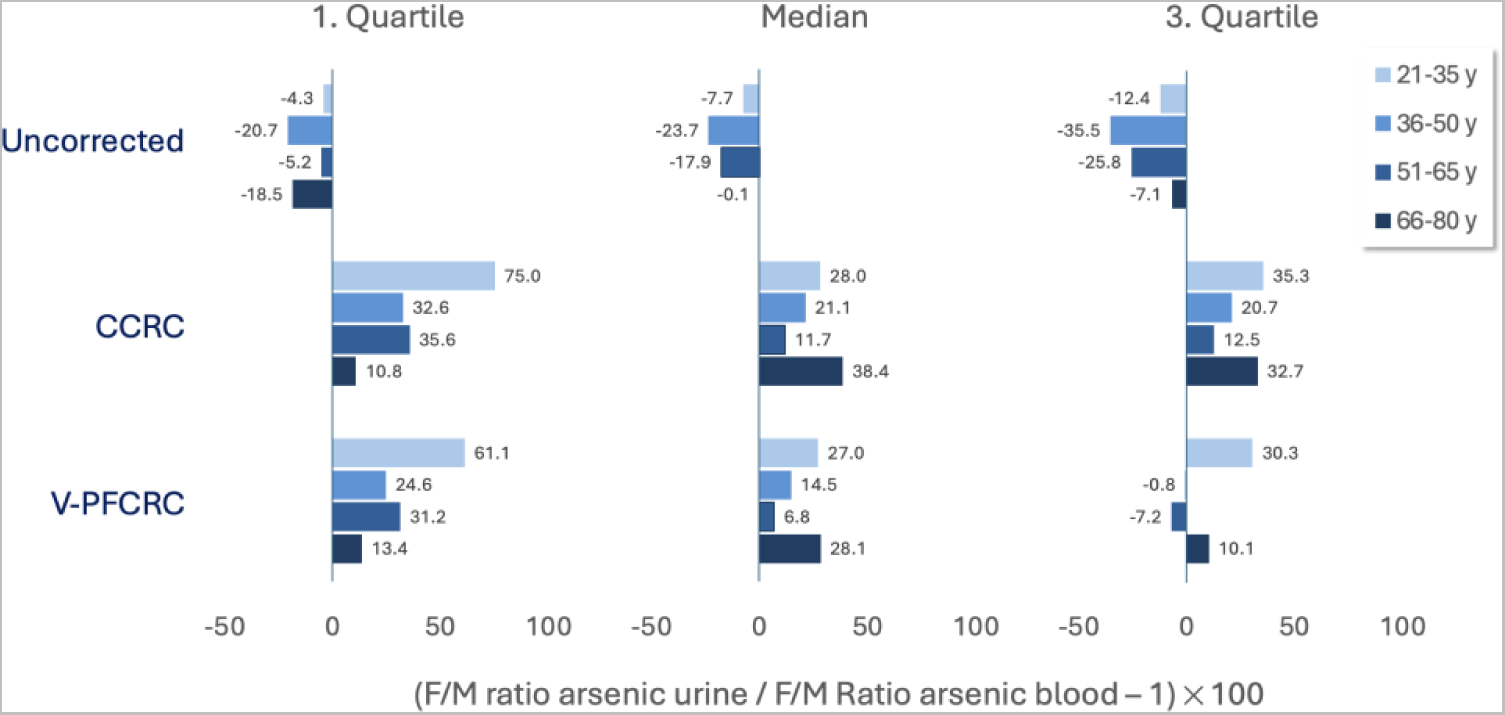
Percentual deviations of urinary from blood sex ratios of Arsenic Quartiles 1-3. The Female/Male ratios of arsenic quartiles 1-3 for urine in UC, CCRC, and V-PFCRC mode were divided by those for blood. The percentual deviation was calculated as shown in the abscissa labeling. Values less than 0 indicate a higher female/male blood ratio than urine and vice versa. The smaller the deviation from zero, the higher the agreement between blood and urine.

It is important to note that while these ratios can indicate the reliability of the dilution adjustment method, differences between blood and urine measurements may not necessarily stem from errors in the measurement process. Instead, they may reflect genuine physiological differences in how males and females metabolize and process arsenic. Potential spurious influences can blur these physiological differences and create a misleadingly better match between blood and urine. This is particularly relevant for uncorrected urine, which generally shows good agreement with blood/sex ratios. This agreement, however, is strongly affected by the gender discrepancies of urinary dilution, artificially widening the gap between female and male arsenic in uncorrected urine and spuriously narrowing it in CCRC mode, as shown in Figure 4. This counteracts any physiological deviations in sex ratios due to more effective female urinary arsenic methylation and excretion. As a result, aligning uncorrected urine with blood may be achieved without accurately reflecting the true efficacy of dilution adjustment.

### 3.5. Effects of Age and Sex on V-PFCRC Equations

To explore the effects of age and sex on the normalization equation, samples were categorized into three age groups. For each group, the log relationship between V-PFCRC normalized arsenic and exponent b was calculated separately for both sexes.

Figure 6 shows the most pronounced differences in curve characteristics observed in the 18-45 age group. Younger men exhibit positive slopes in the log-linear curves, with increasing exponents b for higher arsenic exposures. At the same time, women show substantially negative slopes, with exponents b declining as arsenic levels rise.

**Figure 6:**
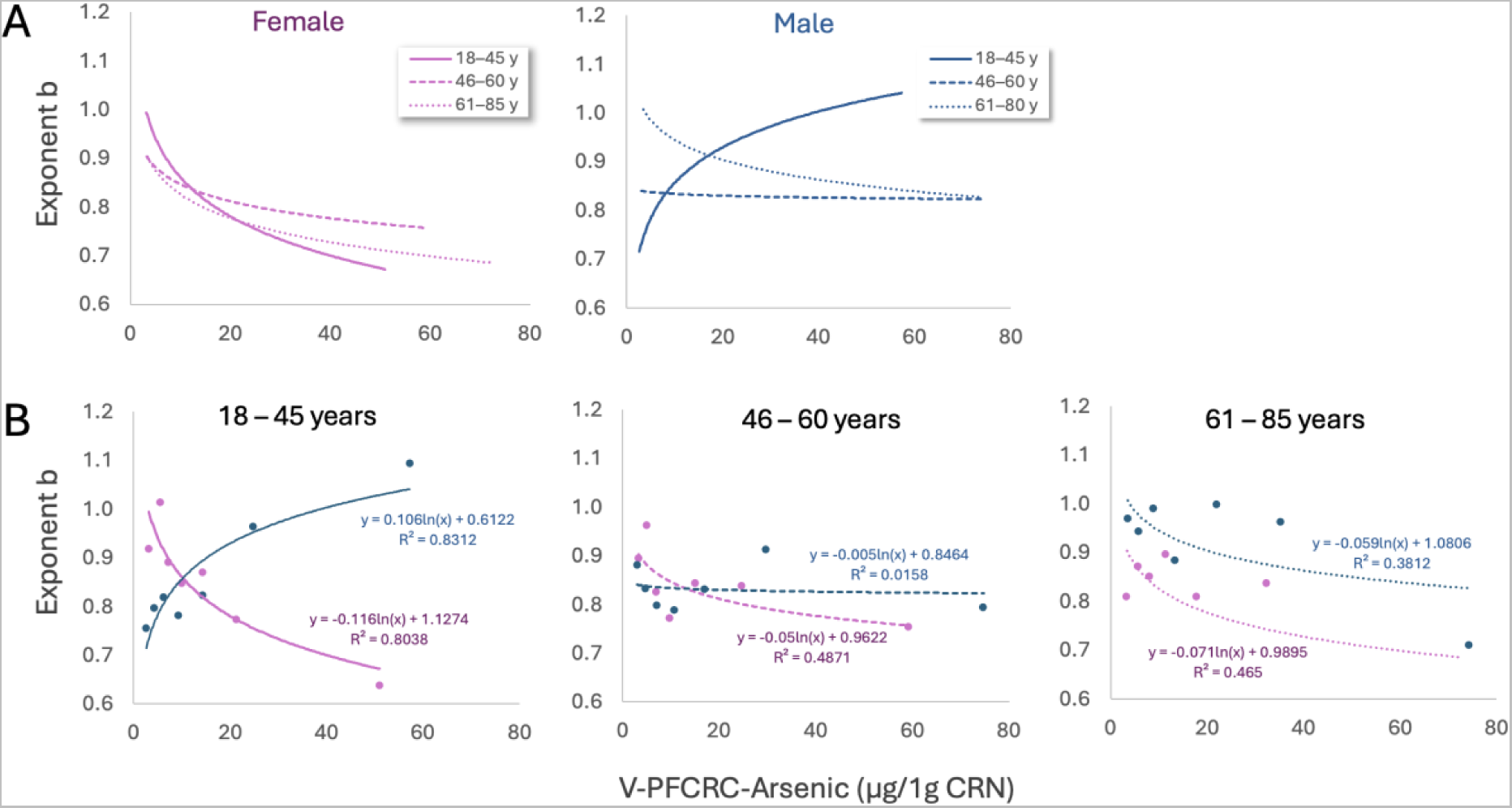
Age- and Sex-differentiated log-linear relationships between Exponents b and V-PFCRC normalized arsenic to 1 g/L CRN. Curves are sorted by either sex (A) or age group (B).

In the middle age group, male b-values experience a significant drop and a flattening of the logarithmic component, marked by a now minimal, negative slope c. In contrast, the female curves for the younger and middle-aged groups differ only slightly, resulting in a broad alignment of the curves for both sexes in middle age.

At older ages, the female curve level decreases while the male curve level rises, both entailing a renewed age difference. However, this difference is much less noticeable in the youngest group. In addition, the curves for both sexes now run mostly parallel due to similar negative slopes, as illustrated in Figure 6b.

### 3.6. Validation of V-PFCRC by Age- and Sex-Stratified Power Functional Regression Analysis

To evaluate the relevance of apparent sex- and age-related differences in V-PFCRC functions for the validity of uniform normalization across various sex- and age-specific subgroups, samples from Set 1 were subdivided into six sex-aggregated age groups and comprehensive sex-aggregated and sex-differentiated cross-age groups. The residual bias between CRN and arsenic was determined for each group using power-functional regression analysis. Arsenic levels in all groups were standardized using the CCRC, V-PFCRC, and S-PFCRC methods outlined in Section 2.5, with identical coefficients and exponents b as detailed in Table 3.

Figure 7 summarizes the slopes and coefficient of determination (R²) for each group’s five urinary result modes. In all subgroups, residual dependence was highest in uncorrected urine, followed by CCRC. All three power-functional result modes performed substantially better than the first two, with only minor differences in both slopes and R² among them. This indicates that applying the sex-differentiated CRN-asymmetry compensated V-PFCRC normalization formulas, determined for sexes between 14 and 82 years,^3^ has significantly mitigated the systemic dilution adjustment error (SDAE) in all studied subgroups.

**Figure 7:**
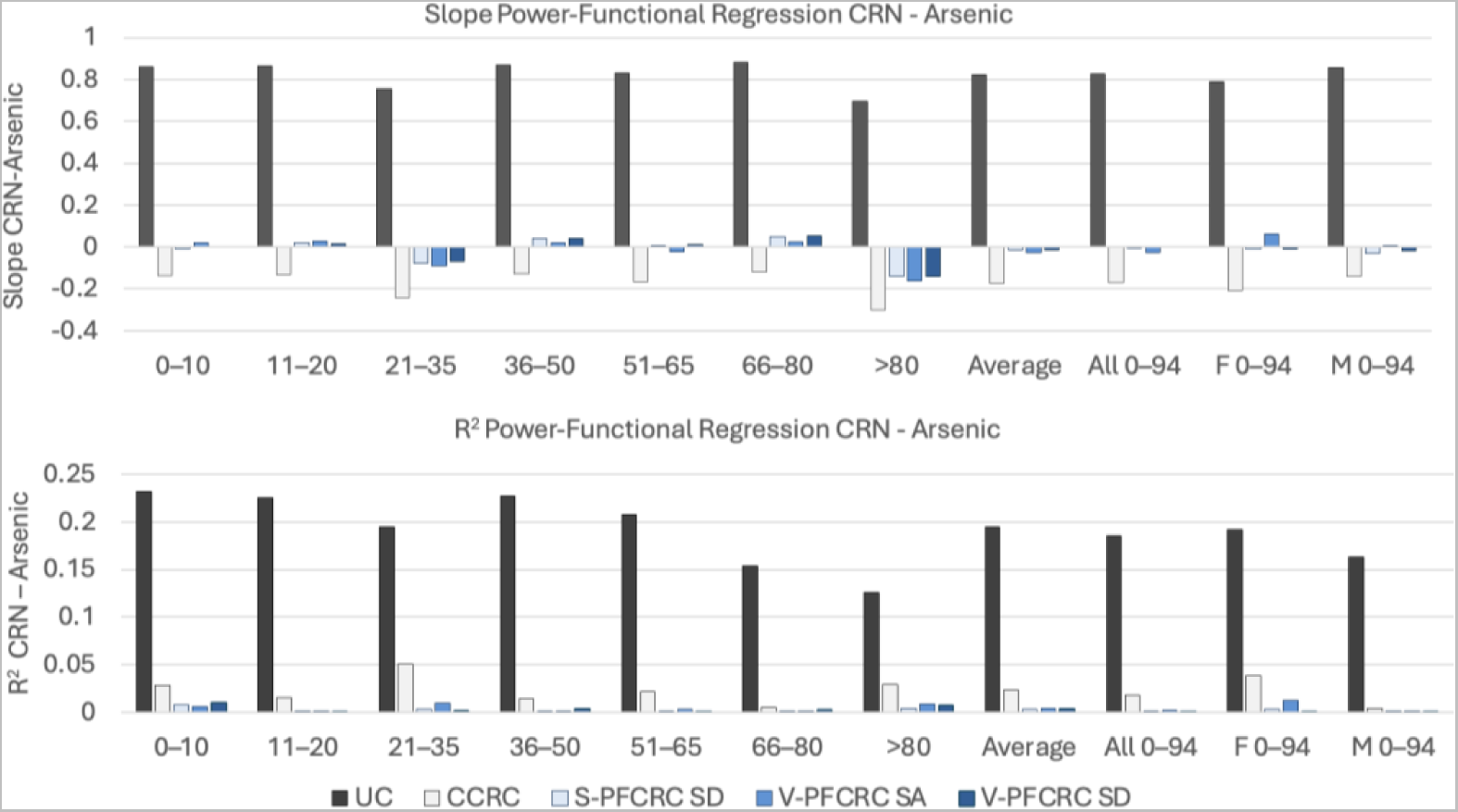
Sex- and age-differentiated efficacy of standardization determined by residual dependence of Arsenic on CRN in various age and sex groups of Set 1. Residual dependence was analyzed based on slopes and R^2^ of power-functional regression analysis of each subset.

Similar power-functional regression analyses were performed for the association of the five urinary arsenic modes with blood arsenic in Set 2. In all subgroups shown in Figure 8, the coefficient of determination (R²) between blood and urinary arsenic improves with CRN correction, whether conventional, simple-, or variable power functional. Following correlation results in entire data sets of previous work,^3^ also the differences in R² values within the CRN corrections are comparatively small and vary between individual subgroups. However, in the overall cross-age analysis, as in previous studies, this analysis also shows a lead for sex-differentiated V-PFCRC SD.

**Figure 8:**
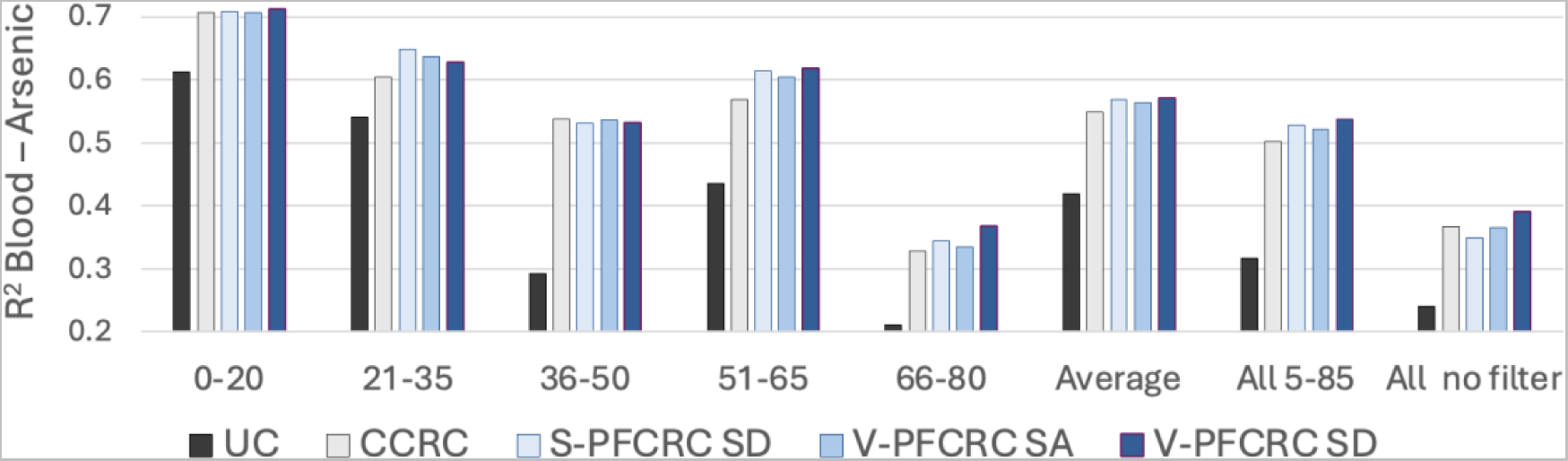
Sex- and age-differentiated associations between arsenic in blood and urine in various subgroups of Set 2. Dependence was analyzed based on R^2^ of power-functional regression analysis of each subset.

Considering the best alignment of sex ratios in Figure 5 for quartile 3 and the previous promising Spearman correlation results with trimmed datasets, excluding values below the median, it is evident that V-PFCRC offers significant advantages over other urinary arsenic result modes, especially at higher and methodologically more reliable blood levels.^3^ As demonstrated previously, This advantage arises from the relatively weak resolution and broad scatter observed in lower blood arsenic levels.^3^

## 4. Discussion and Conclusion

### 4.1. Sex- and Age-related Differences in Arsenic Excretion and V-PFCRC Formulae

The results of this and the previous study strongly suggest that V-PFCRC represents a significant advancement in accurately correcting urinary biomarkers for dilution effects. The method addresses non-linear interactions and biases associated with differential diuresis dependencies through variable power-functional adjustments. The implications of the novel V-PFCRC method extend to various medical and research applications, including the diagnosis and monitoring of diseases, environmental exposure assessments, and forensic investigations. The improved accuracy of urinary biomarker correction can lead to better patient outcomes and more informed public health decisions.

Apart from the significant biases addressed by V-PFCRC, various factors affecting urinary creatinine and arsenic still need to be considered in assessing exposure, even after adjusting for dilution by V-PFCRC. The most critical interconnected factors are lean muscle mass, sex, and age.^50,118,119^ While no direct data was available for lean muscle mass, this study systematically investigates the complex interplay of physiological and biochemical factors influencing urinary arsenic levels, considering age and sex as critical variables. Using an extensive cohort of urinary total weight arsenic samples (Set 1) and a smaller cohort of total weight EDTA blood samples analyzed in parallel (Set 2), the effects of age and sex on arsenic levels across various traditional and power-functional dilution adjustment modes were compared.

The findings reveal significant differences in arsenic excretion patterns in all investigated urinary result modes, blood levels, and V-PFCRC formulae related to age and sex. Estimating the need for further adjustments to these formulae or reference ranges will depend on a deeper understanding of the underlying causes of these differences and their epidemiological significance. Variations in urinary arsenic due to age and sex may result from a range of interconnected factors, summarized in Supplements 1–7. A crucial question is whether the observed sex- and age-related variations in urinary arsenic reflect actual changes in internal or external exposure or if they are driven by age- and sex-related factors affecting the metabolism of arsenic and CRN, fluid regulation, or urine production. After summarizing the key findings of this study, the following section will delve into their intricate biochemical and physiological underpinnings, providing a foundation for further refinement of the V-PFCRC method.

### 4.2. Summary of Key Findings

#### 4.2.1. Impact of Age and Sex on Creatinine Levels

In agreement with existing literature, both sexes exhibited an age-dependent rise in creatinine (CRN) levels, peaking in early adulthood and then continuously declining.^50^ Males generally maintained higher CRN levels than females across most age groups, aligning with the expected physiological differences influenced by muscle mass and hormonal factors.^50,51^

#### 4.2.2. Impact of Age and Sex on Urine Arsenic Levels

Increased urinary arsenic levels were observed with age for both sexes, though the rise in uncorrected arsenic values was more pronounced in males. While CCRC diminished or even reversed this sex difference, it is still noticeable in S-PFCRC and V-PFCRC results and blood arsenic.

#### 4.2.3. Comparison of Urinary and Blood Arsenic Levels

EDTA blood arsenic levels were consistently lower than urinary arsenic levels across all ages, and blood arsenic exhibits marked age increases, especially in males. Age increases in arsenic were more pronounced in blood than in urine, suggesting differential metabolic and excretion dynamics that require further exploration. Sex-differentiated V-PFCRC-SD arsenic concentrations and sex-age dynamics showed more substantial alignment with blood arsenic than conventional CCRC, emphasizing the robustness of V-PFCRC in reflecting accurate arsenic exposure.

#### 4.2.4. Age and Sex-Specific V-PFCRC Equations

Age and sex significantly influenced the log-linear relationships between normalized arsenic and the exponent b, being fundamental to corrective V-PFCRC equations. Younger males exhibited increasing exponents with higher arsenic exposures, while females showed declining exponents. Due to a substantial drop in the male curve level, these differences became less pronounced in middle-aged groups, where both sexes showed more aligned curves. Sex differences re-emerged to a lesser extent in older age groups.

#### 4.2.5. Further Validation of V-PFCRC

The uniform application of the sex-differentiated V-PFCRC formulae, determined initially for a wide age band from 14 to 82 years,^3^ also consistently reduced the residual dependence of arsenic levels on CRN across all age and sex subgroups, demonstrating superior standardization efficacy than conventional correction methods even without further adjustment to sex and age.

Regression analyses of urinary arsenic modes with blood arsenic provided additional evidence of V-PFCRC presenting the most accurate reflection of actual arsenic exposure, supporting its use in diverse demographic settings.

In summary, S-PFCRC and V-PFCRC significantly reduce the substantial CRN bias found in uncorrected urine and the considerable residual CRN bias in CCRC urine. This is demonstrated in Tables 4 and 5 for sets 1 and 2 by the insignificantly low Spearman correlations between S-PFCRC- or V-PFCRC arsenic and CRN).

Although the age factor considerably impacts corrective V-PFCRC formulae, it only has a minor influence on bias removal by V-PFCRC, as clearly shown in the validation results of Figure 7. Broad bias reduction is a universal effect of V-PFCRC that applies to each age group. The differences among age groups are minimal compared to the significant improvements in CRN-bias removal by V-PFCRC from UC or CCRC arsenic.

### 4.3. Physiological, Hormonal, and Toxicokinetic Considerations

#### 4.3.1. Sex Differences in Urine and Blood

The general male increases in urinary concentration in this study indicate a near-synchronous male dominance of CRN and UC arsenic, suggesting extensive commonalities between these two analytes concerning the hydration-dependency of their renal excretory mechanisms. The generally higher male urinary concentration is primarily due to the more substantial impacts of the Renin-Angiotensin-Aldosterone System and Vasopressin (AVP), along with the weaker influence of AVP-attenuating renal prostaglandins and NT-proBNP in men, as summarized in Supplements 1 and 2. Additionally, joint tubular transporters, such as MATE-1, may further synchronize the excretion of CRN and arsenic.^10,14–16,120^

The sex difference in uncorrected urinary arsenic, however, does not seem to be solely caused by general sex differences in urinary dilution, as the analysis of Set 2 revealed a significant age-related increase in male dominance of blood arsenic levels. While CCRC adjustments neutralized the sex difference in arsenic to insignificance in Sets 1 and 2, V-PFCRC in both sets maintained a partly significant male bias, aligning better with the concurrent higher blood arsenic levels in males in Set 2. The lower levels of arsenic found in the blood and urine of females in the older three age groups of Set 2 may indicate reduced exposure to arsenic, possibly due to consuming fewer arsenic-containing foods or absorbing less through the intestines. Alternatively, arsenic exposure and daily weight-adjusted net excretion through urine might be less gender-dissimilar or even female-dominant, with lower urinary arsenic levels in females resulting from their higher daily urine per weight production.^7,9,121^

The gender inverted higher female arsenic medians and upper quartiles in blood and urine among 21-35-year-olds in Set 2 (Figure 4) and 21-30-year-olds in Set 1 (Supplementary Excel File), suggest an influence of sex hormones, which are most abundant in this age group. By contrast, lower arsenic values (quartile 1) do not show this inversion, implying that the effect might be limited to moderate and higher arsenic exposure rather than background levels. As explained in section 1.3, estrogens inhibit the reabsorption of inorganic arsenate in the proximal tubule by downregulating the Na-iPT-II transporter. Additionally, estrogens promote the methylation of inorganic arsenic to DMA, enhancing renal excretion.^34,35^ The enhanced methylation in females facilitates the clearance of arsenic from the liver and other organs into the bloodstream, thereby enhancing the renal excretion of total weight urinary arsenic (TWuAs).^122,123^ Thus, arsenic tissue storage in younger women is expected to be lower than in men, with blood levels comparable or even slightly elevated. With estrogen declining during menopause, these effects weaken, resulting in a male bias in blood and urine arsenic levels in middle and old age, as shown in Figure 4. In this context, the less pronounced late-life urinary arsenic flush of older women in Figures 1 and 2 reflects a smaller female total body burden due to the mentioned methylation advantages, especially at younger, more estrogenic ages.

Adopting sex-differentiate V-PFCRC SD formulas improves dilution adjustments compared to sex-aggregated V-PFCRC SA. These advantages are substantiated by better alignment of sex ratios of urinary to blood arsenic (Figures 1 and 2), significant age differences of V-PFCRC formulas (Figure 6), and the closer agreement to blood in V-PFCRC SD than V-PFCRC SA results in previous research.^3^ When optimizing exposure assessments in intra- and interindividual comparisons, the potential benefits of further modifying sex-differentiated formulas by age are evident based on this analysis. Conversely, when tested on entire collectives with systemic dilution adjustment errors averaging out, the adjustment efficiencies between the two correction modes in Figures 7 and 8 did not reveal any substantial benefits of sex-differentiated V-PFCRC SD over sex-aggregated V-PFCRC SA formulas.

#### 4.3.2. Influence of Age

##### 4.3.2.1 Variations in Arsenic Levels in Urine and Blood

In both data sets and across sexes, a strong positive correlation between CRN and uncorrected arsenic is evident, as shown in Tables 4 and 5). However, age appears to affect these parameters in opposing ways, as illustrated in Figure 1. CRN levels decrease steadily from peak levels in young adulthood, whereas urinary arsenic in Set 1 exhibits a consistent upward trend with age across all result modes and sexes. Set 2 confirms this trend in males, while females show a late-life rise preceded by declines in urinary arsenic. Due to the smaller sample size of Set 2, these observations might be less representative than those in Set 1. Notably, in both sets the age increase of urinary and blood arsenic is consistently more pronounced in men (Figures 1 and 2). This might be due to higher retention of arsenic during younger years and subsequent increased internal exposure from decaying storage tissues in older age.

The systemic computational error from dividing by smaller CRN denominators due to age contributes to the apparent age increments in corrected results. However, this is only one of the factors since increases are seen in uncorrected urine and blood, too. A plausible explanation for elevated blood and urine arsenic in older adults could be higher external and internal exposure. Age-related bone degradation, a long-term storage site for arsenic, may lead to more arsenic being released into circulation,^115,124–126^ while increased intestinal permeability in older adults due to factors such as dysbiosis, medication, and nutrient deficiencies may lead to higher absorption rates of external arsenic, too.^127^

As depicted in Figure 3, arsenic concentrations in blood increase notably with age, exhibiting a more pronounced relative elevation than the rises observed in urine, particularly in the oldest age groups (66-80). This discrepancy suggests that progressive renal insufficiency in older adults, alongside the above-mentioned higher exposure,^4,124–127^ limits the transfer of arsenic from blood to urine, thereby accentuating the increases in blood concentrations relative to urine.

The V-PFCRC formulas also show significant variations with age, particularly in the atypical curve of the younger group of men, as depicted in Figure 6. This indicates a potential to improve the V-PFCRC method using age-stratified correction formulas. Generating and implementing these would be straightforward, provided sufficient large data sets are available. While adopting age-stratified V-PFCRC formulas could yield even more accurate exposure estimates, bias in larger cohorts is not expected to be reduced significantly. Despite more substantial age deviations of the analyte-specific coefficients c and d, as shown in Figure 6, the use of uniform sex-differentiated V-PFCRC formulas generated from data on 14-82-year-olds still substantially mitigated residual biases across the various age groups tested separately (Figure 7). As mentioned, the benefits of age-stratified V-PFCRC formulas over CCRC and S-PFCRC are generally more significant in intra- and interindividual comparisons than in more extensive studies.

##### 4.3.2.2 Role of Common Tubular Transport for Opposing Age Trends of CRN and Arsenic

The inverse age-dependent trends of CRN and urinary arsenic may appear contradictory to a joint transport mechanism initially, but they may support it.^4^ As age advances, lean muscle mass and metabolic turnover decline, reducing the production of nitrogenous waste products like CRN, which are excreted by the kidneys. This reduction in creatinine excretion may lead to enhanced tubular secretion of arsenic in middle-aged and older individuals. This is because common tubular transporters, such as MATE1, may become more available for this purpose, provided the glomerular filtration rate is not significantly impaired, and the tubular excretion of creatinine is not overly strained. The increased methylation capacity with age may further enhance the excretion of total weight arsenic, resulting in higher proportions of more soluble and easily excreted arsenic species.^34,88,128,129^ The increased methylation at higher ages could also be biochemically linked to the declining biosynthesis of creatine and phosphatidylcholine attributable to the lower lean muscle mass. Together, these two compounds require about 80% of the available SAMe, the essential methyl donor in arsenic metabolism.^4^

##### 4.3.2.3 Variation of Urinary Species Proportions with Age

Previous studies have reported significant age-related increases in total arsenic, arsenobetaine, and, to a lesser extent, MMA and DMA, with tendential reductions in inorganic arsenic.^34,39,55,88^ In this study, only total arsenic was measured. However, based on previous findings, it is expected that methylated arsenic components, more abundant in urine, are the main contributors to the age-related rise in arsenic.^39,42,43,88^ A mechanistic link could be the age-increased baseline actions of Arginine Vasopressin (AVP) on the V1-Receptor and the Renin-Angiotensin-Aldosterone-System, both of which inhibit m-KL and enhance tubular reabsorption of PO_4_^−^ and arsenate.^130–133^ Additionally, both hormonal factors limit renal plasma flow and, to a lesser degree, glomerular filtration rate, possibly contributing to increased urinary total weight arsenic in older adults by the mechanisms proposed in Section 4.4.

##### 4.3.2.4 Need for Further Age-Adjustment in Assessment of Arsenic Exposure

Age proved to be a significant confounding factor for urinary arsenic, independent of the dilution adjustment method adopted. Unlike the dilution-based differences between sexes, which sex-differentiated V-PFCRC formulas can better adjust, these methods do not compensate for the impact of age.

The epidemiological parameters, including total body burden or exposure periods represented by urine concentrations of metals and other biomarkers, can vary considerably with age.^115^ While in older individuals, blood and urine levels may broadly reflect an accumulated metal body burden over many years due to redistribution from degraded bone, the levels of the identical biomarker in younger individuals might indicate rather acute exposure fluctuations.^115,124–126^ An interesting example is the age-specific interpretation of urinary cadmium levels. In healthy, younger kidneys, urinary cadmium reflects the total body cadmium burden due to the constant renal cumulation and excretion proportional to renal loads. Impaired kidneys yet excrete higher amounts as cadmium passes from deteriorating tubular cells into the urine. Similar mechanisms may also apply to arsenic stored in bones and kidneys, explaining its age-related increase.^115^

Given these shifts, accurately assessing acute arsenic exposure necessitates age-related adjustments. This applies to both inter-/intra-individual comparisons and cohort studies with asymmetric age distributions. Since the age increases are real, adjustments are essential for all dilution modes, regardless of age-variable V-PFCRC formulas. This can be achieved by integrating age into multilinear covariate models, using age-adjusted reference ranges, or normalizing values to a specific age.^1,34,44^ Notably, adopting age-adjusted V-PFCRC formulas will not adjust for age variation in internal or external exposure. As mentioned earlier, this will only address errors in systemic dilution adjustments more accurately, enhancing the precision of biomarkers, especially in comparisons within and between individuals or studies demonstrating significant variations in urinary dilution and creatinine production among different study groups.

### 4.4. Variations of V-PFCRC Formulae Related to Age and Sex

Figure 6 illustrates the significant sex and age differences in V-PFCRC standardization formulas. These differences are most pronounced in young individuals, while they diminish in middle and older age groups, primarily due to changes in male characteristics. These variations result from the compound influences of multiple sex—and age-specific renal fluid regulatory factors, as detailed in Supplements 1–7.

#### 4.4.1. Pivotal Role of Osmoregulation in Age- and Sex-Variations of V-PFCRC Functions

Evidence in the literature suggests that differences in osmoregulation based on age and sex may be linked to the abovementioned variations. Previous studies have shown that the age-sex dynamics of urinary osmolality exhibit characteristics similar to the relations between V-PFCRC arsenic and exponent b, illustrated in Figure 6.^7^ Young men exhibited a higher estimated urinary osmolality (uOsm) than young women, with these differences diminishing in middle age due to a decrease in male uOsm, as shown in Supplement 14.^7^ This male-dominant difference in uOsm re-emerges in older age matching the reopening gender gap of V-PFCRC curves in that age group. The hypothesized sex and age modifications in osmolality set points and their primary actuators, RAAS (ATII) and AVP (Supplements 1 and 2), likely result in similar alterations in the relationship between V-PFCRC arsenic and exponent b. This concept aligns with mitigating age-related arsenic increases observed following intravenous fluid administration, suppressing AVP/ATII before sample collection (data to be published).

There are parallels between the slopes and exponents b in Figure 6 and the age- and sex-variable renal actions of ATII/AVP summarized in Supplements 1 and 2. The actions of these hormones are generally more pronounced in males, as they are stimulated by androgens and attenuated by estrogen.^7,8,10,17–29^ This corresponds to the higher sex discrepancy in b-values at younger ages. The decline in sex hormone activity towards middle age, more significant in menopausal women, results in gender alignment of AVP/RAAS activities and uOsm. In older age groups, a minor inconsistency between b-values and uOsm for males consists of a tendential drop in uOsm, which does not correspond to the rising trend of b-values.

Conversely, there is good alignment in women as both uOsm and the exponents decrease. This inconsistency in men could be due to different age group distributions in the studies or intrinsic renal aging mechanisms, which are more pronounced in men and cause fluid homeostasis inelasticity, such as declining renal prostaglandin levels and tubular dysfunction.^134–139^ While ATII/AVP activities increase with age, the paradoxical urinary concentrating ability deficit in seniors is likely due to these intrinsic renal causes.^135,140^ In conclusion, the age- and sex-specific variations of exponent b consistently reflect the renal baseline actions of ATII/AVP, influenced by sex hormones in the young and renodegenerative changes in the elderly, irrespective of the age-related decline in renal concentrating capacity.

#### 4.4.2. Mechanistic Explanation of ATII/AVP-Mediated Effects on the Exponent b

As exponent b approaches 1, the mass ratios between arsenic and CRN remain constant across varying hydration/urinary flow rates. Arsenic exponent b-values are typically <1, indicating reduced arsenic/CRN mass ratios at higher urinary concentrations and enhanced ATII/AVP activities.^3^ Therefore, enhanced actions of ATII will either decrease net TWuAs excretion, increase net CRN excretion, or exert any summative effect on both analytes, resulting in relatively higher excretion of CRN than TWuAs in dehydrated states. Increases in b approximate the exponent to 1, indicating better arsenic and CRN secretions balance across the urinary dilution range.

Creatinine and most blood arsenic components are freely glomerularly filtered, so their hydration-dependent excretion discrepancy will be predominantly due to their differential tubular handling.^141–144^ Any relative restriction of tubular function about glomerular filtration rate might thus align the excretion behaviors of CRN and TWuAs, translating into an approximation of b to 1. Age-related tubular renal damage progresses faster in males than in females,^136–139^ which could conclusively explain the reemerging male-dominant age gap in exponents b at higher ages, as illustrated in Figure 6.

Apart from kidney damage, higher baseline ATII/AVP activities in males and at higher ages can limit tubular relative to glomerular function.^139,145,146^ Angiotensin II (ATII) preferentially constricts efferent arterioles, increasing glomerular filtration pressure but reducing renal and peritubular capillary flow, thus impeding tubular function. Arginine Vasopressin (AVP), primarily involved in water reabsorption in the collecting ducts, also contributes to renal vasoconstriction. Together, these hormones can exacerbate tubular hypoxia and injury, enhancing glomerular at the expense of tubular function.^70,147,148^

The effects of ATII and AVP on CRN excretion are twofold. In prolonged or excessive ATII/AVP activity, such as during advanced volume loss or dehydration, GFR decreases, leading to increased tubular CRN secretion.^111,146^ Conversely, moderate ATII/AVP activity, which is more common, primarily increases the glomerular capillary hydrostatic pressure (GCHP) and enhances GFR, reducing the tubular contribution to urinary CRN.^70,149,150^ This could explain the generally increased exponent b in men, as their higher baseline ATII/AVP activities may enhance GFR, reduce the relative contribution of the tubules to CRN excretion, and thus counterbalance discrepancies in the tubular handling of arsenic and CRN.

The tubular excretion of arsenic species also depends on urinary flow rate and corresponding ATII/AVP activity yet differs from CRN in this regard. Although there is a significant knowledge gap concerning the tubular handling of different arsenic species, it is known that at least the reabsorption of inorganic arsenate (iAsV) is favored by slower urinary flow.^4,151^ More dehydrated states, associated with higher ATII and AVP activities, result in higher tubular reabsorption and lower net secretion of arsenate.

In such states, the increased excretion of creatinine (CRN) leads to a shift in the mass ratio between CRN and arsenic in favor of CRN. This shift in more dehydrated states with elevated ATII/AVP activities can plausibly explain why the exponents of arsenic generally have values smaller than 1.

Contradicting this explanation is the association of higher inorganic arsenate (iAs_V_) reabsorption with higher methylation and total weight arsenic, which is the relevant parameter for this study.^34,38,39,55^ The methylation process of arsenate and its increasing effect on TWuAs may lag the reabsorption of arsenate and, therefore, not immediately affect urinary TWuAs.

The restraining effects of prolonged ATII/AVP elevation on membrane protein Klotho (m-KL) expression further complicate these mechanisms. Lower m-KL levels increase arsenate (iAsV) reabsorption and favor its methylation, leading to higher proportions of easily excreted methylated TWuAs components and increased TWuAs, as outlined in Sections 1.4 and 1.5.

There is empirical evidence of opposing ATII/AVP effects on net tubular excretion of iAs (inhibitory) and TWuAs (stimulatory), which can explain lower %iAs and higher TWuAs in seniors and gender-identical TWuAs excretion in males despite female methylation advantages.^4,34^ Males could thus compensate for methylation drawbacks with enhanced TWuAs excretion-promoting effects of ATII/AVP.

In summary, prolonged states of lower m-KL expression associated with higher ATII/AVP activities, such as in males and older age, generally seem to increase exponents b and thus reduce the systemic dilution correction error (SDAE).

The increasing exponents b in young men with rising arsenic levels, contrasted by opposite trends in all other groups, require additional explanation. This might be due to the high CRN production in young men, which may cause faster saturations of tubular CRN transporters in dehydrated states, shifting TWuAs/CRN mass ratios in dehydration towards arsenic, entailing higher exponents b. Higher arsenic exposure could intensify this effect through additional competition and impairment of common tubular transporters, explaining the positive slope of coefficient c as detailed in Section 1.1 and preliminary work.^3^

Additionally, the sex-differentiated methylation capacity of arsenic could play a role. Interestingly, at lower arsenic levels, which in the present study population generally can be assumed to have higher proportions of inorganic arsenic, the exponent b of young women is higher than that of men and much closer to 1. The female methylation advantage could be noticeable here, as higher proportions of methylated arsenic can be better excreted in urine and behave more similarly to CRN in the tubule. This effect diminishes at higher arsenic levels, where mostly organic loads with predominantly methylated arsenic are present. At higher arsenic exposures, the advantage of female tubular CRN secretion over arsenobetaine in the dehydrated state may become more significant, causing a relatively steep negative slope (coefficient c) in young women.

These theoretical considerations rationalize sex-age-specific differences in the corrective formulas (Figure 6). Yet further research is needed to validate these findings and to elucidate the tubular handling of the individual arsenic species.^4,151^

### 4.5. Validation, Advantages, and Limitations of V-PFCRC

The apparent age and gender differences in the V-PFCRC relations only marginally compromise the standardization quality within the individual subgroups, as shown in Figures 7 and 8. This corroborates the significantly improved precision and comparability of V-PFCRC results across all age groups and sexes compared to uncorrected and CCRC arsenic. The improvements of sex-differentiated V-PFCRC SD over alternative power functional corrections tested in this study (sex aggregated V-PFCRC SA, sex-differentiated simple S-PFCRC) were relatively narrow. The benefits of V-PFCRC SD are more evident in comparisons within separate analyte levels rather than in entire collectives tested in this study. This is attributed to adequate compensation of non-linear, exposure-differentiated skews, including complex metabolic and excretory interactions between analytes and CRN. By correctly assuming power-functional rather than linear relationships between uAs_UC_ and CRN, a significant source of error in dilution adjustment in extensive cohort studies can be neutralized despite remaining biases unaccounted for by V-PFCRC. Compared to uncorrected and CCRC values, simple power-functional corrections have significantly improved correlations of standardized total urinary arsenic (TuAs) with drinking water exposures.^2^

While the advantages of V-PFCRC over CCRC and S-PFCRC are evident, how does it compare with more elaborated covariate-adjusted standardizations, which reportedly improved results over classical models? In general, the V-PFCRC method corrects a substantial bias brought about by erroneously assumed linearity in factually non-linear physiological relations. In covariate-adjusted models, this notable misconception is either symptomatically addressed by restricting CRN ranges or ignored within accepted limits.^1,3,34,152^ This confers on V-PFCRC the advantage of dispensing with CRN range restrictions and associated sample rejects. At the same time, more reliable, dilution-independent standardized values are achieved in intraindividual and, with minor limitations, interindividual comparisons. Fewer sample rejects imply resource savings and the disposal of more valid samples, increasing the power of any epidemiological study involving urinary biomarkers.

Despite the mathematical adjustment of this significant bias by V-PFCRC, residual inaccuracies may result from the unaccounted impact of other biasing factors on CRN and the analyte, primarily age and lean muscle mass (BMI) and, to a lesser extent, comorbidities, ethnicity, exercise, nutrition, and the time of sample collection.^1,34,152^ The combined spurious influence of these biases on CRN production, secretion, and any CRN correction will average out in comparable cohorts with sufficient subject numbers. Various factors can also influence CRN in opposite directions, thus moderating the summative distorting effect on corrected results, particularly in larger cohorts. The results in this study on arsenic and iodine in the previous study demonstrated better correlations of all conventional and power-functional CRN-corrected arsenic and iodine modes with EDTA blood (arsenic) or serum (iodine).^3^ There is evidence that cumulative biases on CRN production might be less relevant for accuracy than the systemic dilution adjustment error addressed in this paper and previous work.^2^ They may still cause considerable misinterpretation in exposure analyses of smaller groups and comparative studies of dissimilar individuals or collectives concerning covariates influencing creatine formation and CRN. In pursuing optimized precision, it can be worthwhile to accommodate these factors additionally.

While a sex-differentiated, age-adjusted variable power-functional correction (V-PFCRC) or a similar adjustment by SG or uOsm is expected to yield sufficiently reliable results for most clinical, forensic, and epidemiological scenarios, thereby reducing sample rejections, it may be necessary to use more complex covariate-adjusted models or even integrate. V-PFCRC into them, depending on the study design. A promising future approach could synthesize advanced covariate-adjusted standardization plus creatinine covariate adjustment with the V-PFCRC presented in this study.^1,34^ Ideally, three sequential standardization steps should be adopted in a more sophisticated correction procedure:

1. Standardization of CRN (SG, or uOsm) for age, BMI, sex, and other factors by multiple regression covariate adjustment.
2. Calculating covariate-CRN adjusted V-PFCRC values for analytes (here, all TWuAs species separately) using V-PFCRC formulas determined with covariate-adjusted CRN.
3. Standardization of the obtained V-PFCRC values for age (e.g., 50 years).

### 4.5. Conclusions

The V-PFCRC method significantly advances accurately correcting urinary biomarkers for dilution effects. This method effectively addresses non-linear interactions and biases associated with differential diuresis dependencies by incorporating variable power-functional adjustments. In clinical and epidemiological contexts, adopting V-PFCRC can lead to more reliable assessments of urinary biomarkers, ultimately improving the diagnosis, monitoring, and research of various medical conditions and environmental exposures.

The findings confirm the significant improvements of the novel V-PFCRC method over uncorrected and conventionally CRN-corrected urine in rigorous age- and sex-stratified analysis and, in comparison with blood arsenic detected in parallel.

Based on age-sex-stratified formula generation and literature review, sex-age-kinetics of P-VFCRC corrective formulae were shown to align well with urinary osmolality, predominantly determined by the activities of the Renin-Angiotensin-Aldosterone System (RAAS) and Arginine Vasopressin (AVP).

While the novel sex-differentiated V-PFCRC method largely accounted for sex biases, the presented results underscore the potential for further refinement of V-PFCRC formulae according to age, as it proved to significantly affect both arsenic exposure and the most common default dilution correctors, such as CRN, osmolality, and specific gravity.^2,3^

Future research should expand on the physiological and biochemical underpinnings of these differences, mainly focusing on hormonal influences and metabolic pathways. Moreover, validation across broader populations and with additional biomarkers will enhance the accuracy, generalizability, and applicability of the V-PFCRC method.

## Data Availability

All data produced in the present study are available upon reasonable request to the authors

## Abbreviations

ATII: Angiotensin II
AVP: Arginine vasopressin (= Adiuretin, ADH)
CCRC: Conventional Creatinine Correction
CKD: Chronic Kidney Disease
CRN: Creatinine
DMA: Dimethylarsinic acid
E: Estrogen
FGF23: Fibroblast Growth Factor 23
GCHP: Glomerular Capillary Hydrostatic Pressure
GFR: Glomerular Filtration Rate
iAs: Inorganic Arsenic (iAs_III_ + iAs_V_)
%iAs: Percentage of iAs in Total weight arsenic
iAs_III_: Trivalent Arsenite
iAs_V_: Pentavalent Arsenate
ICP-MS: Inductively Coupled Plasma Mass Spectrometry
MATE1: Multidrug and Toxin Extrusion Protein 1
m-KL: Proteohormone transmembrane Klotho
MMA: Monomethylarsonic acid
NaPi-IIa: Sodium-coupled PO_4_^−^ Co-Transporters
NT-proBNP: N-terminal pro B-type Natriuretic Peptide
oAs: Organic Arsenic (Sum of DMA + MMA, Arsenobetaine, Arseno–sugars, –lipids, and –choline)
OCT2: Organic Cation Transporter 2
uOsm: Urinary Osmolality
P: Progesterone
PO_4_^−^: Phosphate
RAAS: Renin-Angiotensin-Aldosterone System
RPF: Renal Plasma Flow
SAMe: S-Adenosyl-Methionine
S-PFCRC: Simple Power Functional Creatinine Correction using fixed exponents
T: Testosterone
TuAs: Sum of iAs and methylated primary metabolites of iAs (iAs + DMA + MMA)
TWuAs: Total Weight Urinary Arsenic = all ICP-MS detectable forms of inorganic and organic arsenic (measured in this study)
As_UC_: Uncorrected TWuAs in µg/L
uAs_C_: Conventionally Creatinine-corrected TWuAs
uAs_N_: Urinary Total Weight Arsenic, normalized to 1 g/L CRN by V-PFCRC SD
UC: Uncorrected urinary result mode (in µg/L for Arsenic)
uOsm: Urinary Osmolality
V-PFCRC: Variable Power Functional Creatinine Correction (all forms)
V-PFCRC SA: Sex-Aggregated Variable Power Functional Creatinine Correction using identical analyte-specific coefficients c and d for both sexes
V-PFCRC SD: Sex-Differentiated Variable Power Functional Creatinine Correction using sex-specific coefficients c and d

## 5. Acknowledgements

Dr. Katrin Huesker, department manager of Special Immunology, IMD Berlin, is thanked for providing anonymized spot urine data.

## 6. Author contribution

TCC conceptualized and designed the work, reviewed the literature, prepared, evaluated, interpreted, and illustrated the data, developed the corrective S-PFCRC and V-PFCRC formulae, drafted, substantially revised, and submitted the paper

## 8. Ethics

The study was exempt from ethical approval by the Ethics Committee of Northwest and Central Switzerland (EKNZ), 4056 Basel, based on retrospective analysis of anonymized data without clinical information other than urinary creatinine/TWuAs, age in years, sex, and exam date. The purpose was methodological quality improvement only. There was no pharmacological or other medical intervention, risk for physical or psychological harm, identifiability, or breach of confidentiality for any subject involved.

## 11. Supplementary Material

**Supplement 1:**
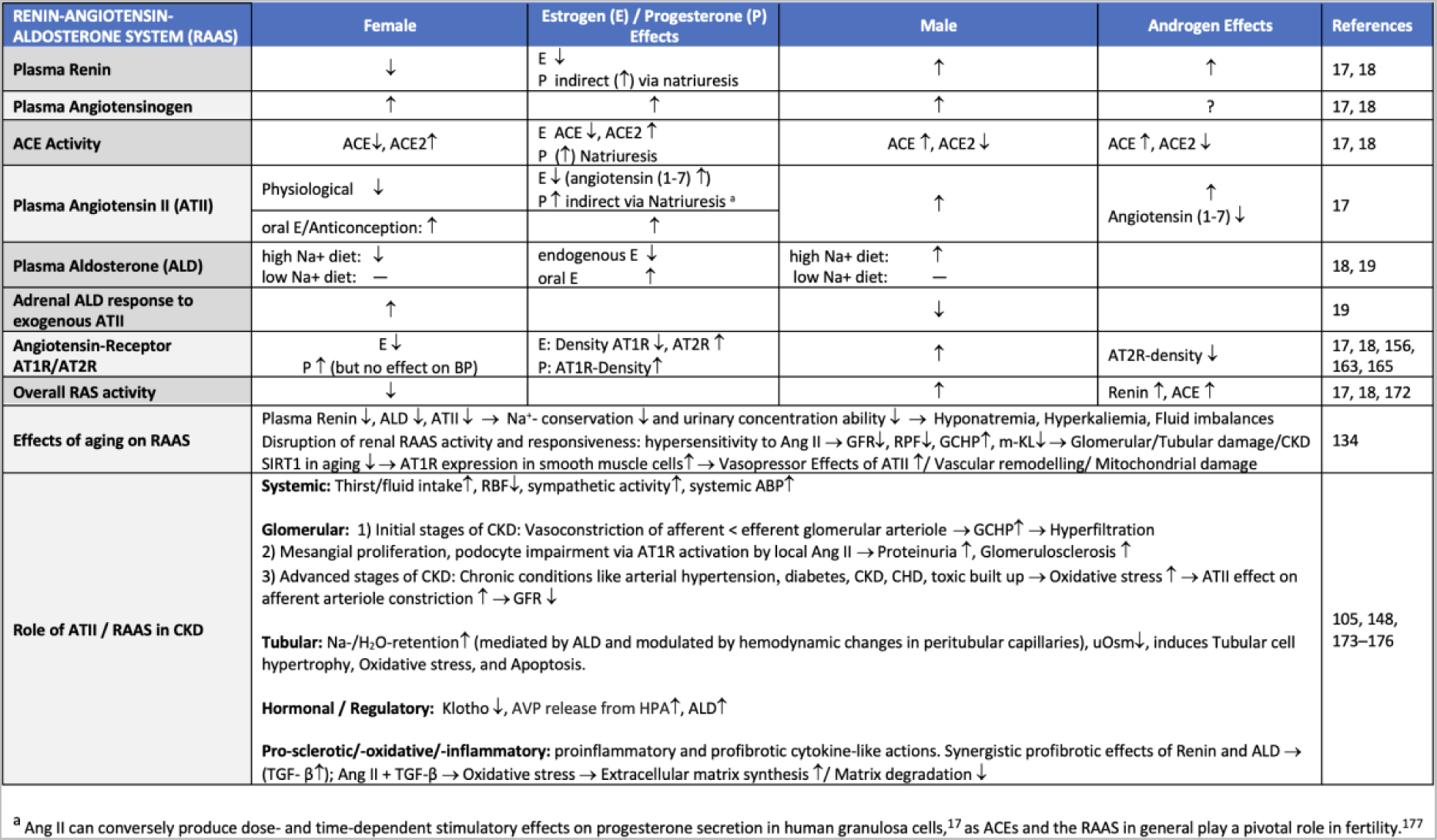
Sex-differential hormonal regulation of osmo- and fluid balance by the Renin-Angiotensin-Aldosterone-System (RAAS).

**Supplement 2:**
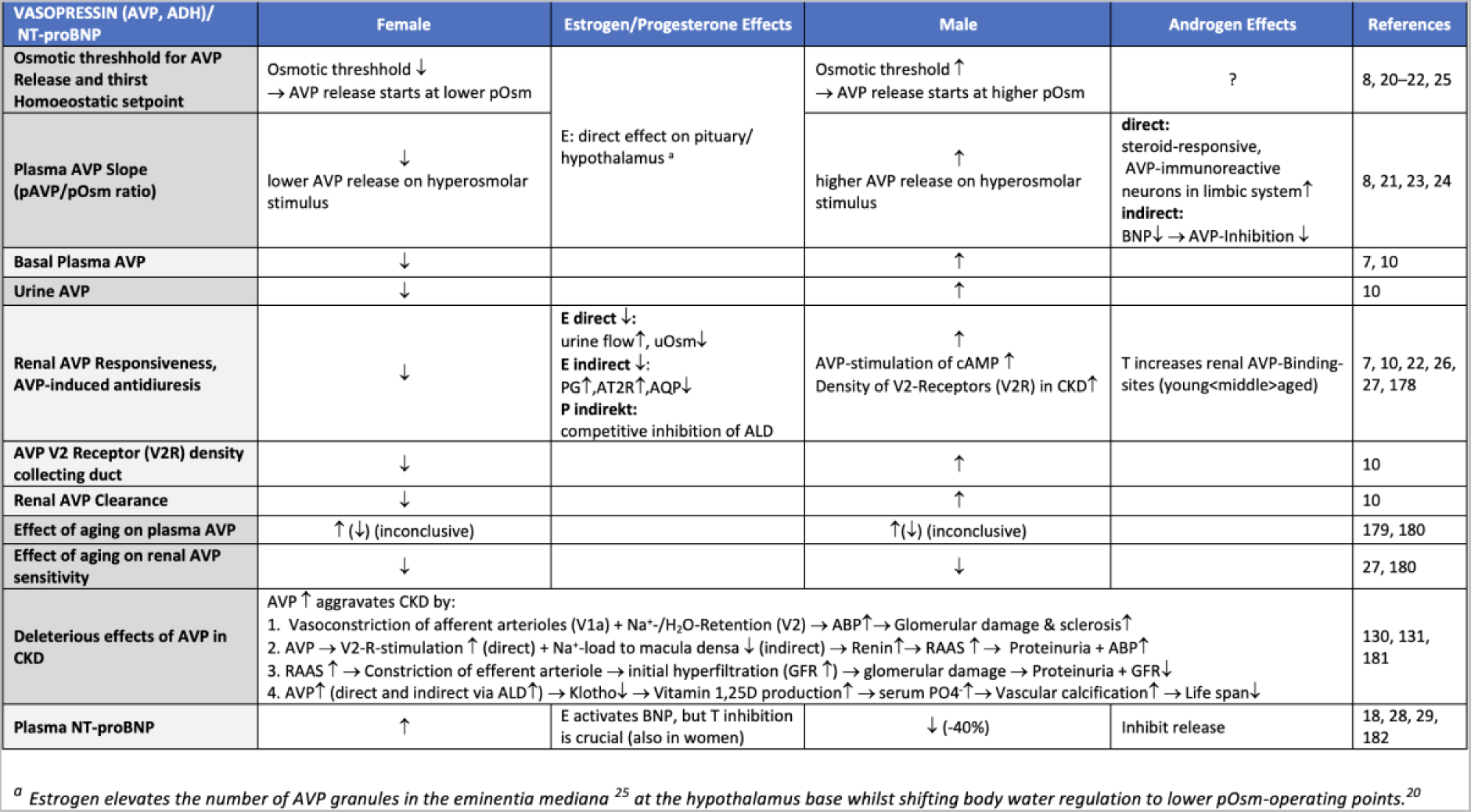
Sex-specific hormonal regulation of osmotic and fluid balance by Arginine Vasopressin (AVP) / Antidiuretic Hormone (ADH) and its antagonist N-terminal pro-B-type Natriuretic Peptide (NT-proBNP).

**Supplement 3:**
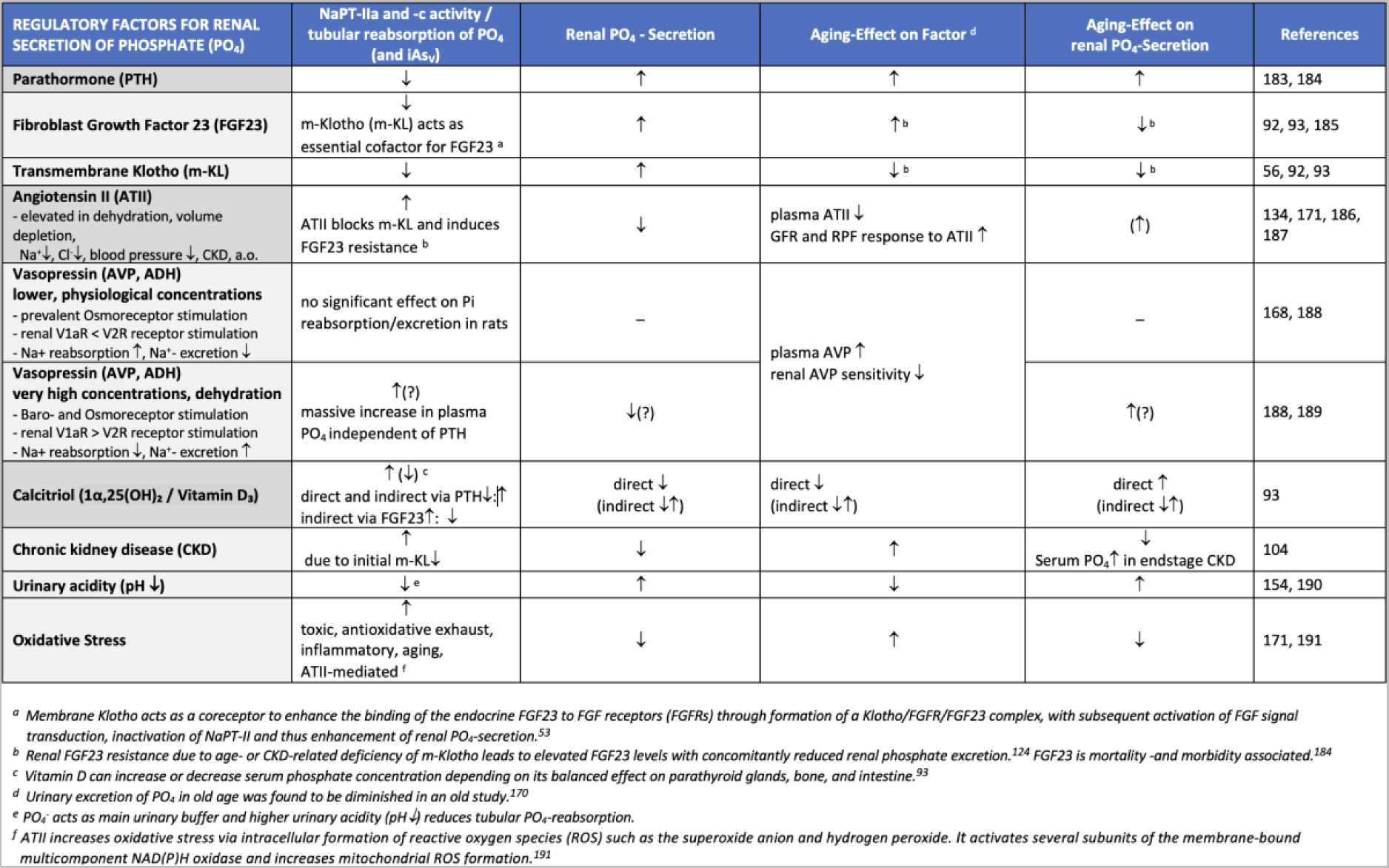
Effects of aging on hormonal and pathological factors affecting renal excretion of Phosphate (PO_4_^−^) and inorganic Arsenate (iAs).

**Supplement 4:** Sex-specific influence of sex hormones on renal membrane transporters.

Supplement 3: Effects of aging on hormonal and pathological factors affecting renal excretion of Phosphate (PO<sub>4</sub><sup>-</sup>) and inorganic Arsenate (iAs).
| RENAL TRANSPORTERS | Female | Estrogen/Progesterone Effects | Male | Androgen Effects | References |
| --- | --- | --- | --- | --- | --- |
| Pool size of transporters in proximal tubules (PT) | ↓<br>(NaPi2, NHE3, cldn2, a.o.) | direct / indirect | ↑<br>(NaPi2, NHE3, cldn2, a.o.) | direct / indirect | 10 |
| Pool size of transporters in distal tubules (DT) | ↑<br>(NCC, NKCC2, SPAK, AQP2, cldn7) | direct / indirect | ↓<br>(NCC, NKCC2, SPAK, AQP2, cldn7) | direct / indirect | 10 |
| Upregulation of OATs and OCTs (Organic Anion- and Cation Transporters)<br>- mainly in S3-segment of PT | OAT2, OAT5, SGLT1, MRP3 | OAT2, OAT5 | OAT1, OAT3, OCT2, OATP1, OATP1a1, OATP3a1, MRP4, MATE1, PG1/MDR1, MDR1a, URAT1, BCRP, CLDN-1, -2, -11, -16 | OAT1, OAT3, OCT2, OATP1a1, OATP3a1, URAT1, MRP4, MATE1 | 10, 160 |
| Downregulation of OATs and OCTs<br>- mainly in S3-segment of PT | OAT1, OAT3, OCT2, OATP1, URAT1, MATE1, MRP4, PGy1/MDR1, MDR1a, OATP1a1, OATP3a1, BCRP, CLDN-1, -2, -11, -16 | OAT1, OAT3, OCT2, BCRP, MATE1 | OAT2, OAT5, SGLT1 | OAT2, OAT5, SGLT1 | 10 |
| Aquaporins (AQP)<br>- only AQP2 is responsive to AVP | AQP1↓, AQ2↓, AQP3↑ | AQP1↓, AQ2↓, AQP3↑ | AQP1↑, AQ2↑, AQ3↓ | AQ3↓ | 10, 161, 162 |
| Sex Hormone receptors (humans) | ERβ (IST, CD, PT)<br>PgR B (Glo, DT, IST)<br>AR (DT) ↓ |  | ERα (IST, CD)<br>ERβ (IST, CD, PT)<br>PgR B (Glo, DT, IST)<br>AR (DT) ↑ |  | 10 |

**Supplement 5:** Physiological sex differences and influence of sex hormones on basic urinary parameters.

Supplement 4: Sex-specific influence of sex hormones on renal membrane transporters.
| URINE PARAMETERS | Female | Estrogen/Progesterone Effects | Male | Androgen Effects | References |
| --- | --- | --- | --- | --- | --- |
| 24h Urine volume | Inconsistent (↑) |  | in 4/9 studies: 10-14% ↓<br>in 5/9 studies: ± 5%, not significant |  | 7 |
| Urine concentration | ↓ | indirect | ↑ | indirect | 7 |
| Water turnover / -excretion | ↑ |  | ↓ |  | 8 |
| Natriuresis (Sodium excretion) | ↑ | P: direct (Prox. & Dist. Tubule) ↓<br>P: indirect (ALD-Antagonist) ↑<br>E: ↓ | ↓ | 5α-DHT → ATII mediated NHE3↑<br>T → ENaC mRNA↑<br>T → ADH-effects ↑<br>→ reabsorption Na<sup>+</sup>, HCO<sub>3</sub>, fluid↑ | 10, 30, 31 |
| Urine osmolality | ↓ | indirect | ↑ (+21-39%) | indirect | 7, 10, 11 |
| Urine osmolality - effect of aging | decline > 50 years |  | decline > 40 years |  | 7 |
| Urinary excreted total osmotic load | ↓ |  | ↑ |  | 7 |
| Urine Creatinine | ↓ | indirect:<br>lean muscle ↓<br>OCT2 ↓<br>MATE ↓ | ↑ | direct stimulation<br>indirect:<br>lean muscle ↑<br>ADH ↑, OCT2 ↑, MATE1 ↑ | 7, 10, 11, 12 |
| Urine pH<br>→ pH ↓ with aging | ↑<br>Crucial GI anion uptake ↑ |  | ↓ |  | 153 |
| Tubular H<sup>+</sup>, Cl<sup>-</sup> and HCO<sub>3</sub><sup>-</sup> - Transport | AE1/Band 3, NHE1, NHE2, Pendrin ↑ | E: Cytosolic carbonic anhydrase ↑ | CLDN-1, -2, -11, -16 ↑ | Cytosolic carbonic anhydrase ↓ | 10, 153, 154, 155 |

**Supplement 6:**
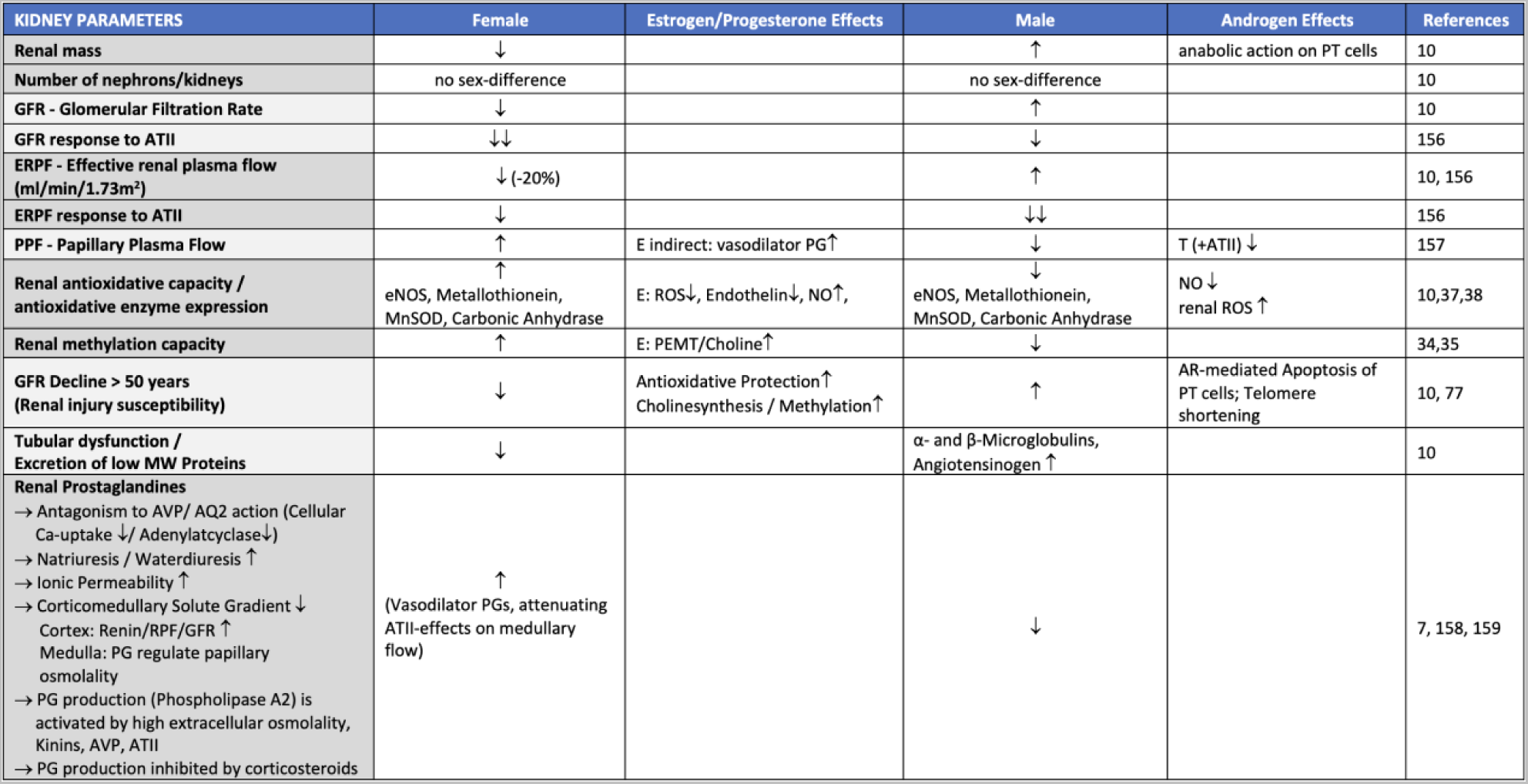
Influence of sex hormones on basic renal parameters.

**Supplement 7:** Influence of sex hormones on systemic fluid distribution.

Supplement 6: Influence of sex hormones on basic renal parameters.
| SYSTEMIC PARAMETERS | Female | Estrogen/Progesterone Effects | Male | Androgen Effects | References |
| --- | --- | --- | --- | --- | --- |
| Blood Pressure | Physiological and transdermal E2:<br>↓ | E Vasodilatation, NO↑, AT2R↑<br>P Na+-Excretion ↑ via<br>ALD-Antagonism <sup>a</sup> | ↑ | permissive on ATII<br>AT1R/AT2R Ratio↑ | 17, 19, 163,<br>164, 165 |
|  | High dose/oral E2, Gestagens:<br>↑ | plasma ALD↑ |  |  |  |
| NO/Vasodilation | ↑ | E ↑ crosstalk with MasR<br>P ↑↓ (situative) | ↓ | NO ↓ | 10, 17, 37 |
| Extracellular Volume | ↓ (F < M) | E/P direct ↑<br>E indirect (RAS↑, ADH↑) | ↑ | direct ↑: Na<sup>+</sup> - retention at proximal tubules<br>indirect ↑:<br>- permissive effect on HGH-induced Na<sup>+</sup> - retention<br>- BNP↓ → AVP-Inhibition ↓ | 19, 20, 32,<br>33 |
| Plasma Volume | 65 ml/kg | E ↑ (TER-Albumin ↓)<br>P ↓ | 70 ml/kg | ? | 20, 32 |
| Serum Phosphate (PO<sub>4</sub>, sP) | Progressive decline until 45 years | E ↓, NaPT-IIa ↓, Ca-reabsorption↑ | Progressive decline until 45 years | T ↓ (?) | 53, 98, 104,<br>166–171 |
|  | <45 years: no sex difference<br>>45 years: ↑ |  | <45 years: no sex difference<br>>45 years: ↓ |  |  |
| Whole body metabolic rate | ↓ |  | ↑ |  | 10 |
| Nitrogenous waste products (Creatinine, Urea, Uric acid) | ↓ |  | ↑ |  | 10 |

<sup>a</sup> Despite potentially elevating Ang II and AT1R levels, progesterone does not appear to raise blood pressure in response to ATII in women.<sup>40</sup>

**Supplement 8:**
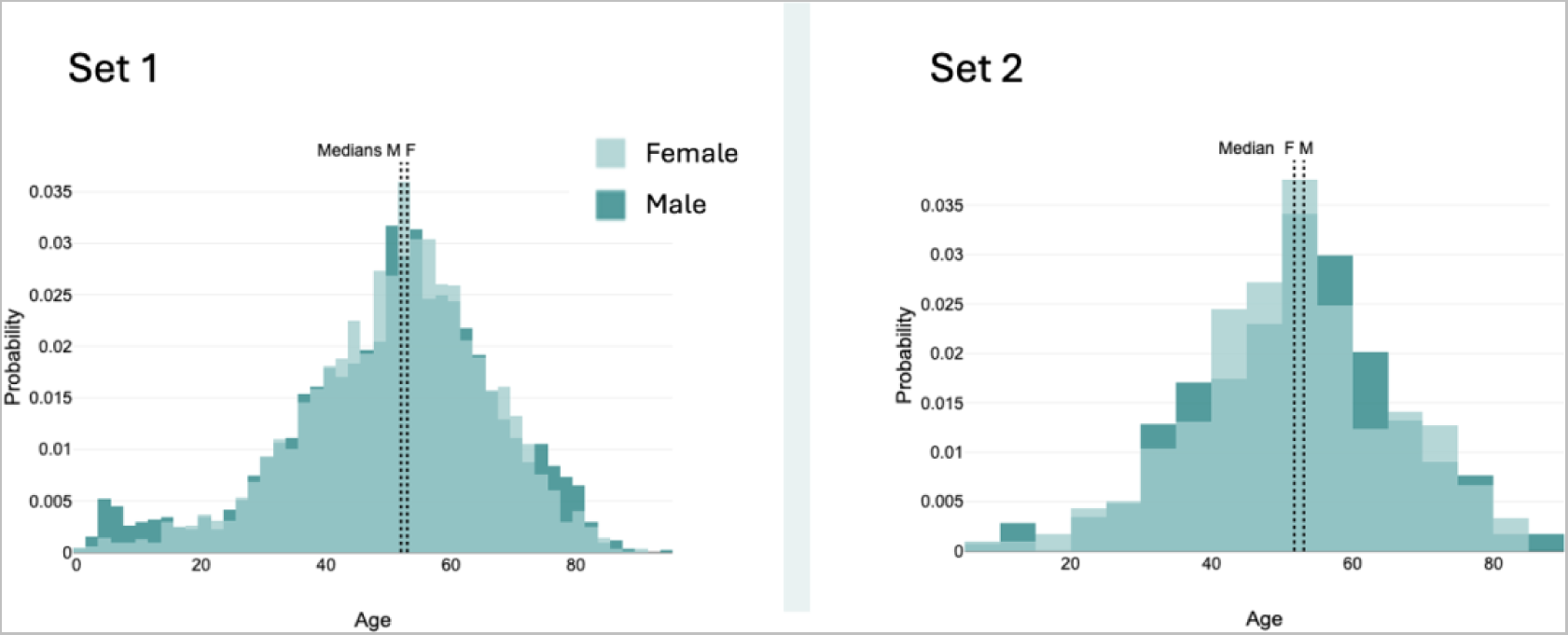
Sex-specific age distribution of sets 1 and 2. Age is presented in years as linear data.

**Supplement 9:**
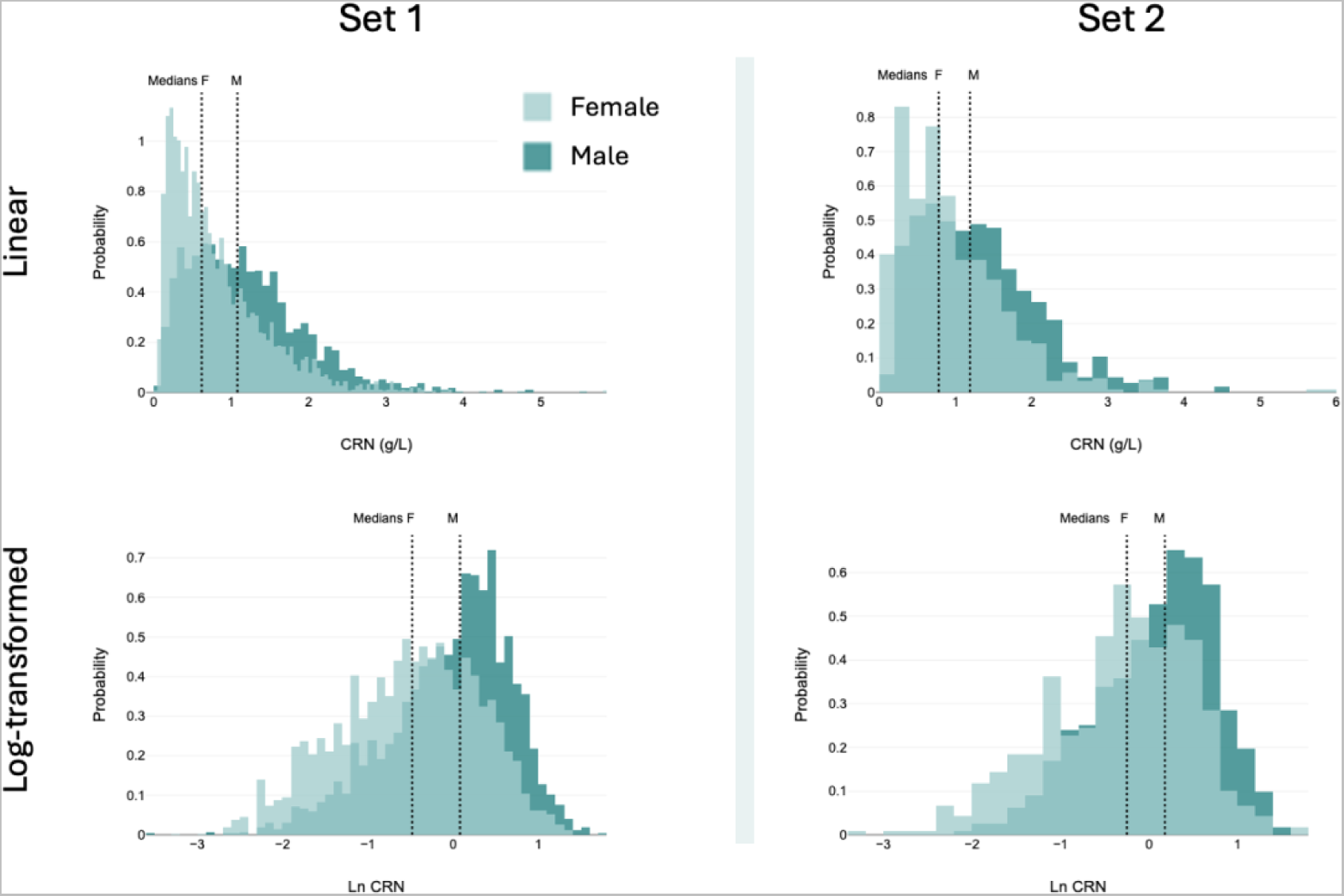
Sex-specific CRN distribution of sets 1 and 2. CRN data in g/L is presented in linear and log-transformed form. Linear data are left skewed, log data are right skewed yet closer to normal distribution.

**Supplement 10:**
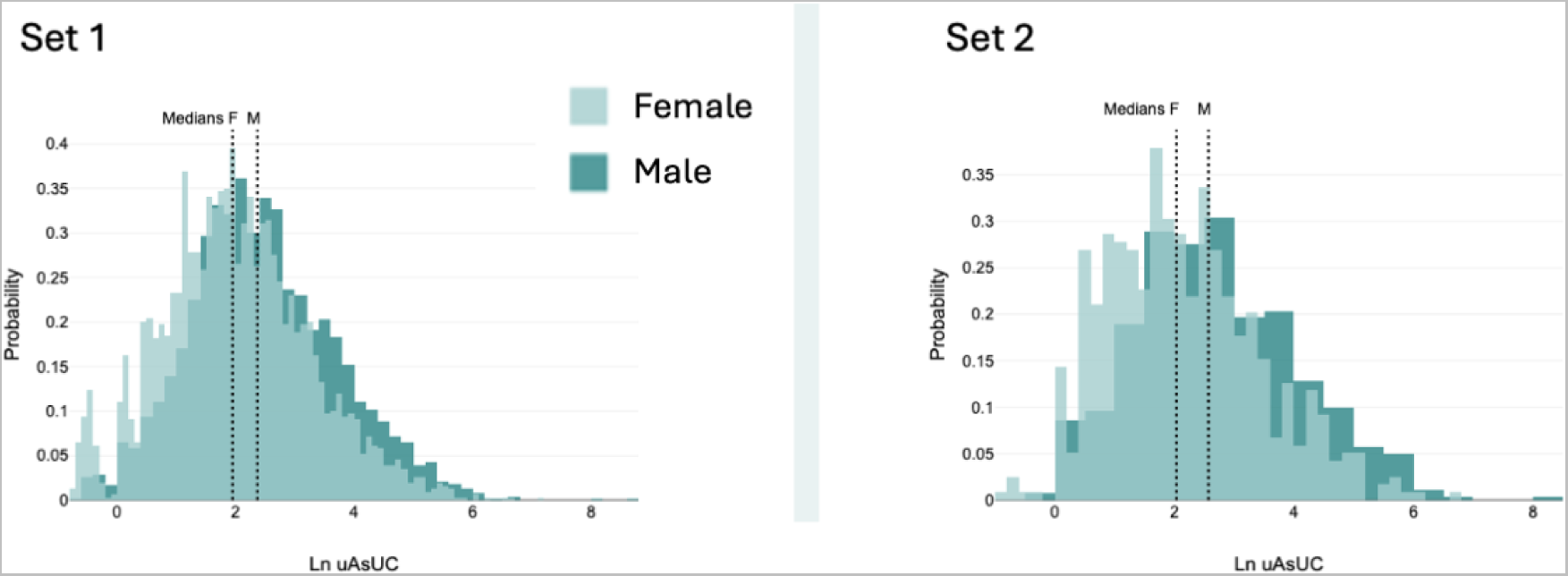
Sex-specific uncorrected urinary arsenic (uAs_UC_) distribution of sets 1 and 2. Uncorrected Arsenic in µg/L is presented in log-transformed form. Linear data are strongly left-skewed, while log data show a near-normal distribution in both sets.

**Supplement 11:**
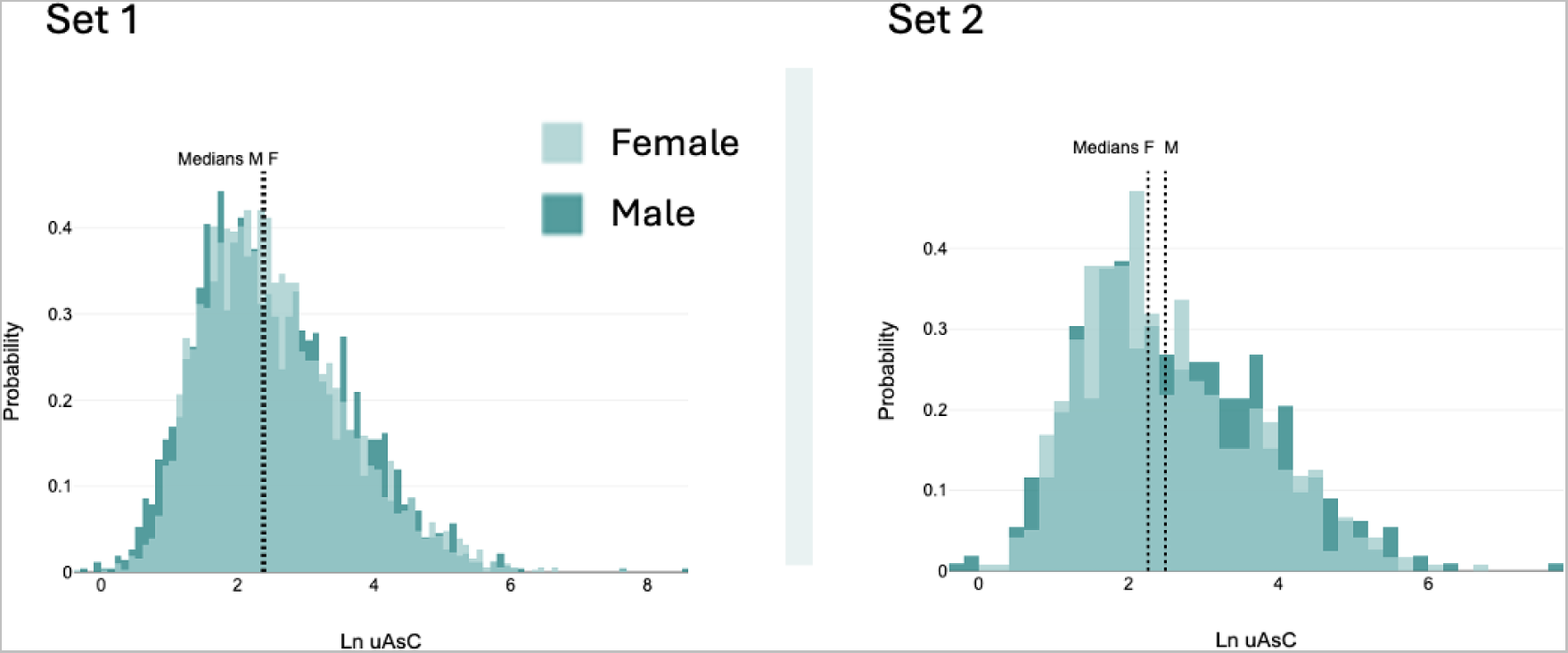
Sex-specific CCRC arsenic (uAs_C_) distribution of sets 1 and 2. CCRC Arsenic in µg/g CRN in log-transformed form. Linear data are strongly left-skewed, while log data show a less pronounced left skew in both sets.

**Supplement 12:**
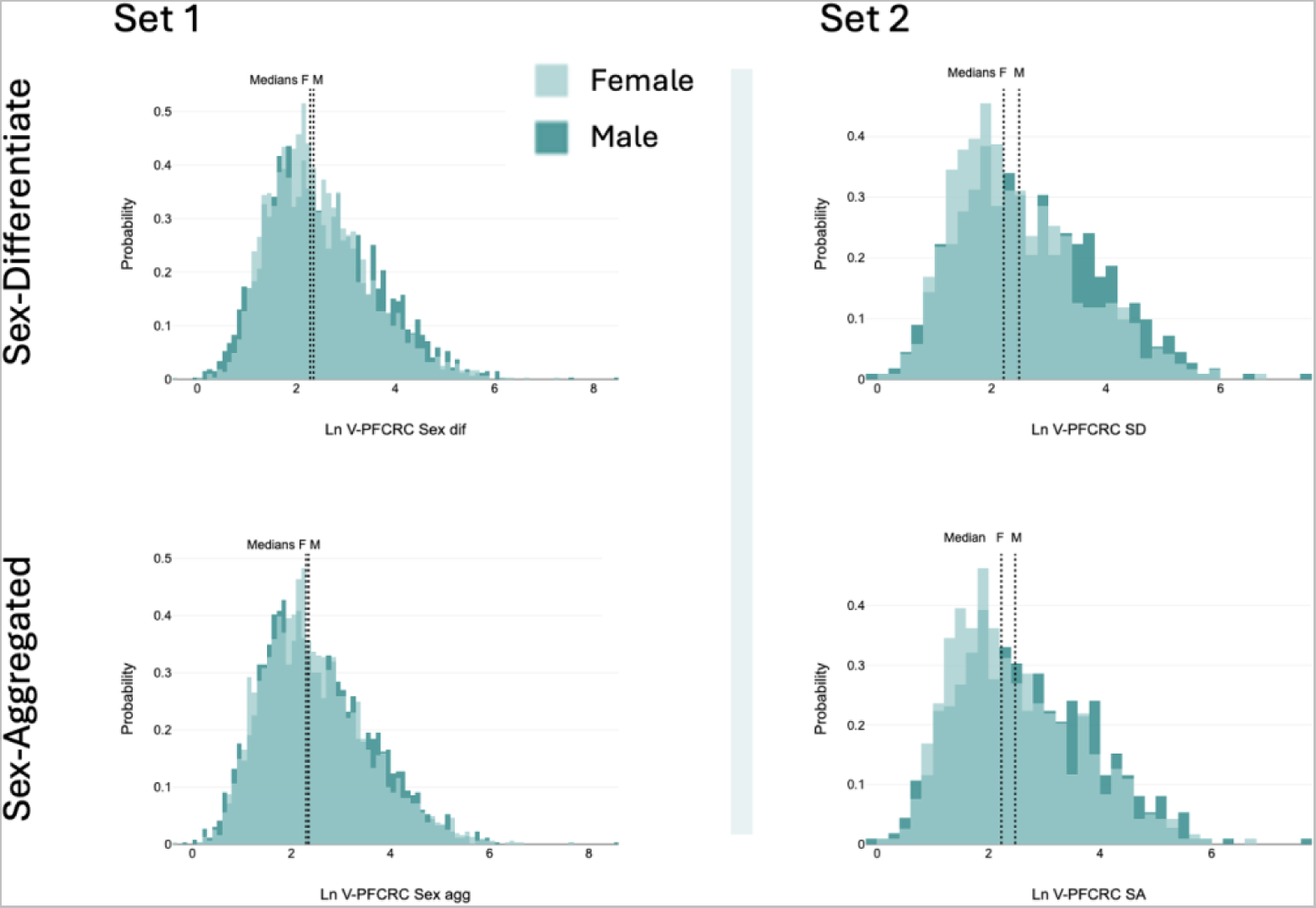
Sex-specific V-PFCRC arsenic (uAs_N_) distribution of sets 1 and 2. V-PFCRC Arsenic in µg/1g CRN in log-transformed form is given for sex-aggregated and -differentiated corrective formulae. Linear data are strongly left-skewed, while log data of both sets are less pronounced left-skewed in both sets.

**Supplement 13:**
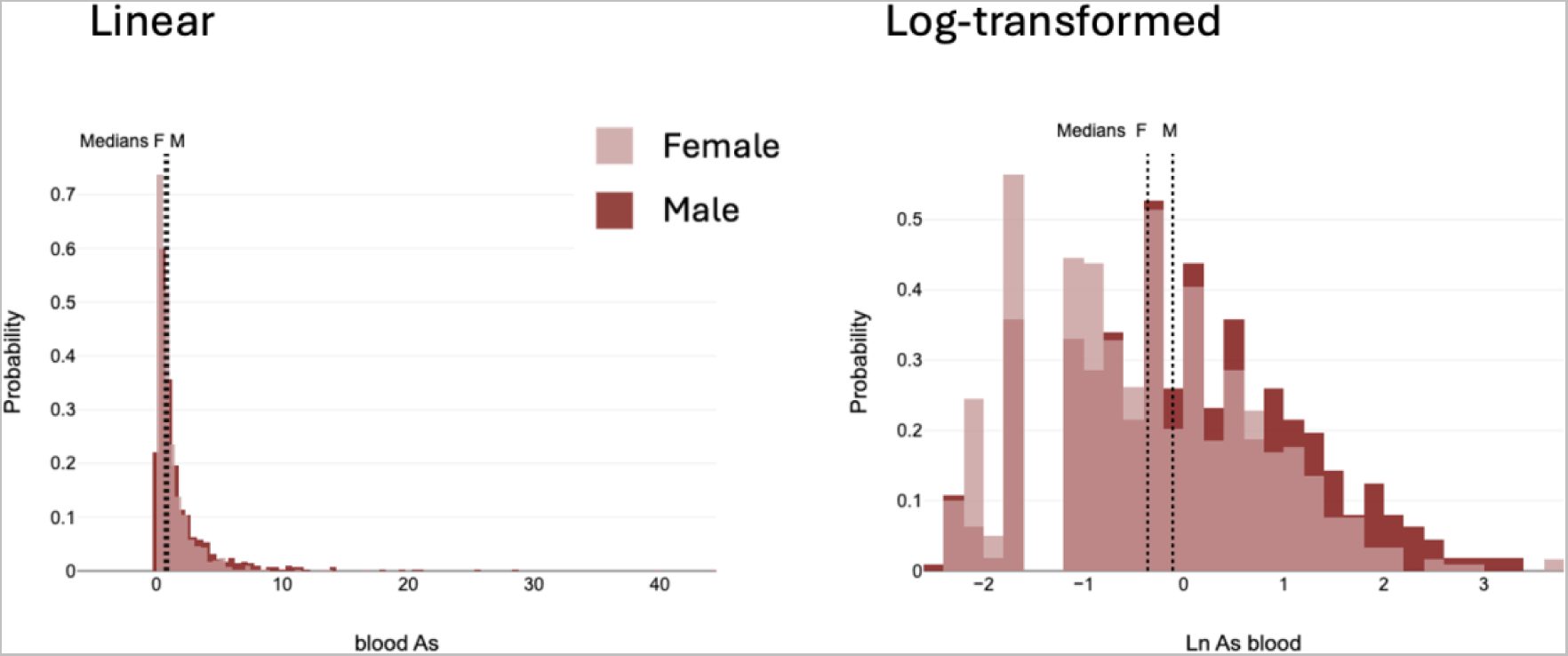
Sex-specific blood arsenic distribution of sets 2. Blood Arsenic in µg/L in linear and log-transformed mode. Linear data are strongly left-skewed, while log data of both sexes are nearly normally distributed.

**Supplement 14:**
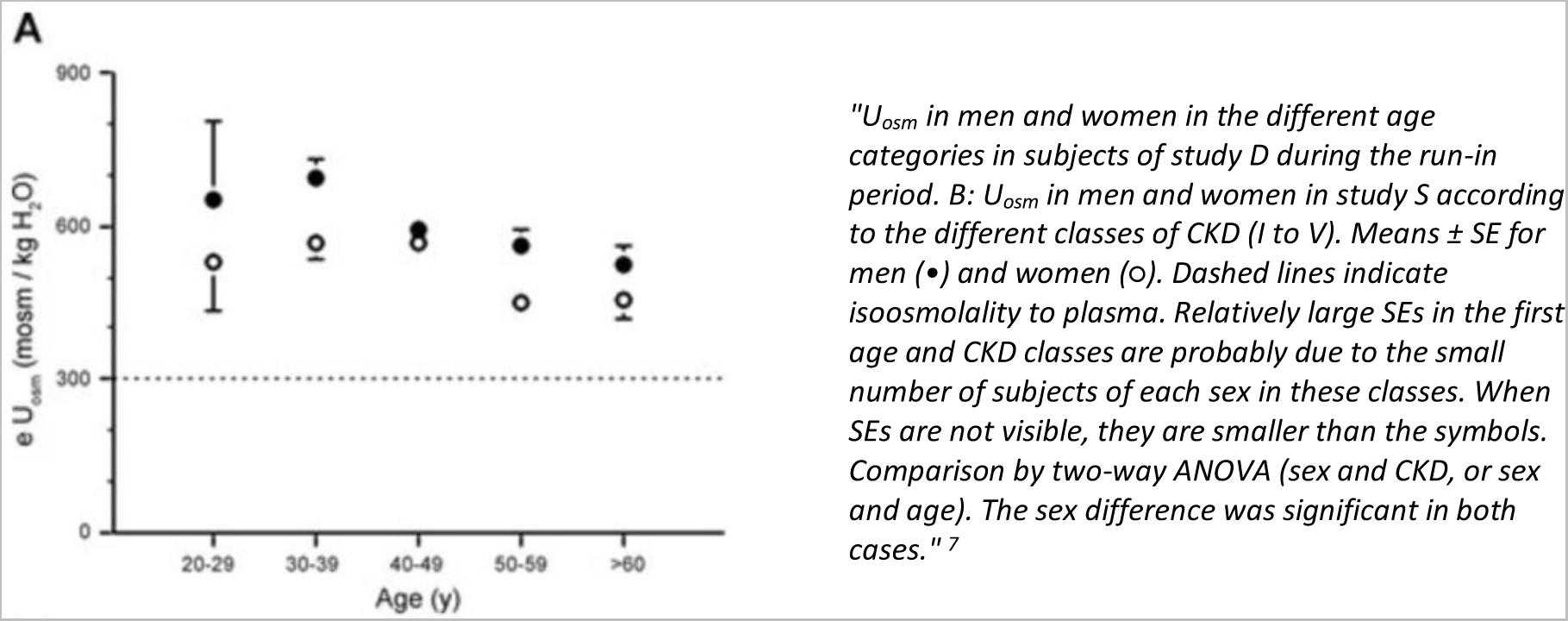
Effect of age on sex-differentiated urine osmolality (Perucca J et al., 2006). Reproduction of original Figure 2 A of “Sex difference in urine concentration across differing ages, sodium intake, and level of kidney disease,” Perucca J, et al. (2006).^7^

